# Neuromodulation with Ultrasound: Hypotheses on the Directionality of Effects and Community Resource

**DOI:** 10.1101/2024.06.14.24308829

**Authors:** Hugo Caffaratti, Ben Slater, Nour Shaheen, Ariane Rhone, Ryan Calmus, Michael Kritikos, Sukhbinder Kumar, Brian Dlouhy, Hiroyuki Oya, Tim Griffiths, Aaron D. Boes, Nicholas Trapp, Marcus Kaiser, Jérôme Sallet, Matthew I. Banks, Matthew A. Howard, Mario Zanaty, Christopher I. Petkov

## Abstract

Low-intensity Transcranial Ultrasound Stimulation is a promising non-invasive technique for brain stimulation and focal neuromodulation. Research with humans and animal models has raised the possibility that TUS can be biased towards enhancing or suppressing neural function. Here, we first collate a set of hypotheses on the directionality of TUS effects and conduct an initial meta-analysis on available healthy human participant TUS studies reporting stimulation parameters and outcomes (*n =* 47 studies, 52 experiments). In initial exploratory analyses with univariate tests, we find that parameters such as the intensity and continuity of stimulation (duty cycle) show statistical trends towards likely TUS neural enhancement or suppression of function. Machine-learning analyses were limited by the currently small sample size. Given that human TUS sample sizes are expected to increase, predictability on the directionality of TUS effects could improve if a database is available. Therefore, we establish an *inTUS* database and resource for the systematic reporting of TUS parameters and outcomes to assist in greater precision in TUS use for brain stimulation and neuromodulation. The paper concludes with a selective review of human clinical TUS studies illustrating how hypotheses on the directionality of TUS effects could be developed for empirical testing in the intended clinical application, not limited to the examples provided.

**Highlights:**

- Collated set of hypotheses on using TUS to bias towards neural enhancement or suppression
- Meta-analysis results identify parameters that may bias directionality of TUS effects
- *inTUS* resource established for systematic reporting of TUS parameters and outcomes
- Selective review of patient TUS studies for enhancing or suppressing neural function

## INTRODUCTION

In the last decade, low-intensity focused Transcranial Ultrasound Stimulation (TUS) has emerged as a promising non-invasive brain stimulation technique for neuromodulation in research and clinical settings. TUS uses sound waves—in the sub-MHz range—that pass through the skull to deliver focal acoustic energy onto a targeted brain area. Compared to other more established non-invasive brain stimulation techniques, such as Transcranial Magnetic Stimulation (TMS), transcranial Direct Current Stimulation (tDCS) or transcranial Alternating Current Stimulation (tACS), TUS offers several advantages: i) focal deep brain targeting (Fig. 1); ii) multi-target, including bi-hemispheric, stimulation capabilities; and, iii) neuromodulatory effects that can last tens of milliseconds to hours after the sonication period has ended (Blackmore et al., 2023; Deffieux et al., 2015; Deffieux et al., 2013; Legon et al., 2014; Mueller et al., 2014). The neural effects of TUS depend on factors including the intensity and duration of the stimulating acoustic wave. In this review, we primarily focus on *low-intensity* TUS as used for neuromodulation (typically <50 W/cm^2)^; (Lee et al., 2021; Murphy et al., 2024) with some consideration of *moderate-intensity applications* (>190 W/cm^2^) used for perturbing the blood-brain barrier (Kim et al., 2021; Spivak et al., 2022; T. Zhang et al., 2021) and *high-intensity focused ultrasound* (up to 10,000 W/cm^2^) used for clinical thermal ablation in neurosurgery patients (Zhou, 2011). The duration of TUS stimulation and its effects is a key factor, with immediate effects during TUS stimulation referred to as ‘online’ effects and those that can last after TUS stimulation referred to as ‘offline’ effects.

**Figure 1.**
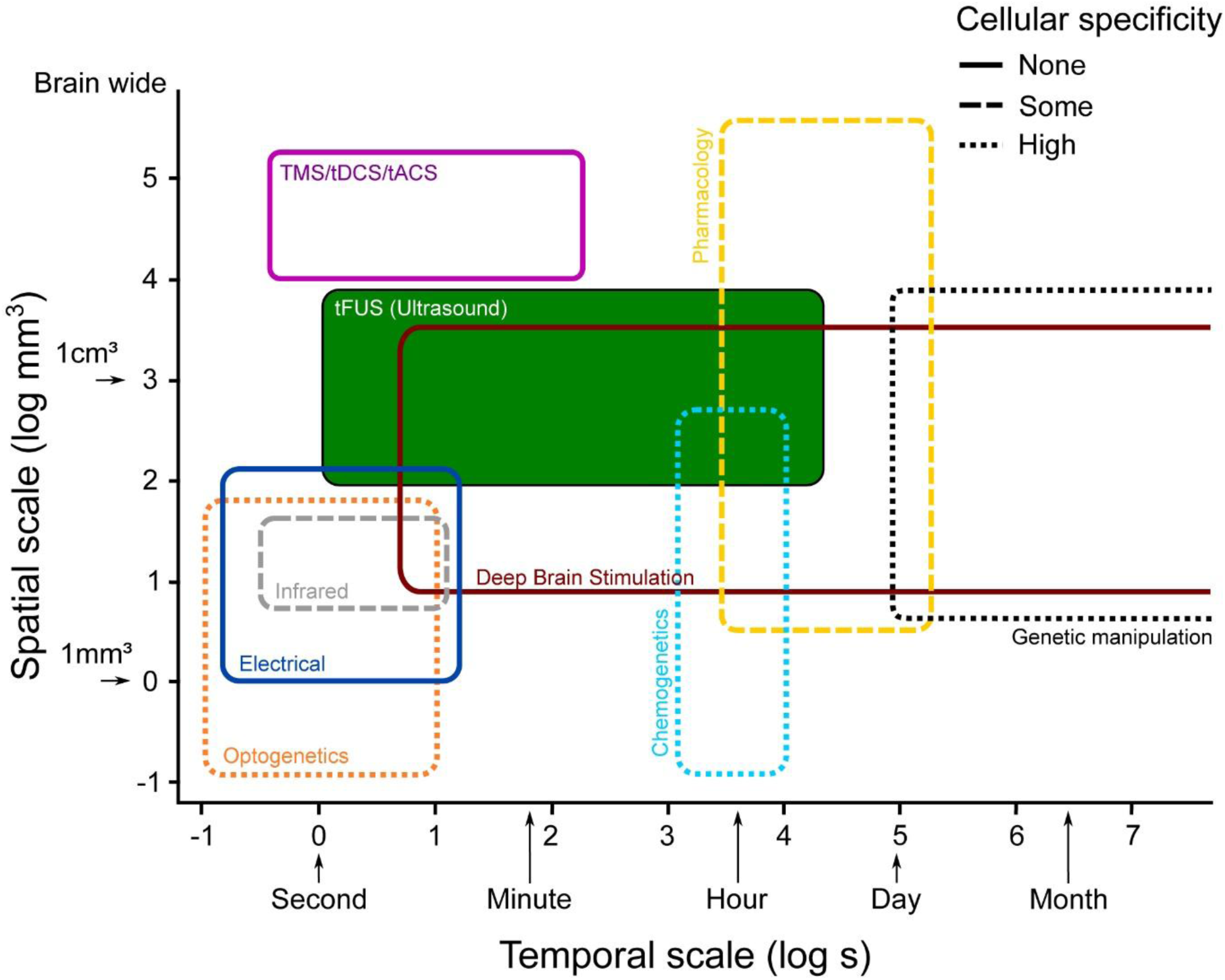
Schematic overview of the specificity of brain perturbation techniques. Brain perturbation techniques vary in the precision of the spatial and temporal effects that can be elicited, on logarithmic (log10) scales. This includes transcranial Focused Ultrasound Stimulation (TUS), in green. Some approaches with cellular specificity are shown that are currently primarily in use with nonhuman animals as models (optogenetics, infrared neuromodulation, chemogenetics and genetic manipulation). Figure modified with permission from P.C. Klink, from (Klink et al., 2021).

**Figure 2.**
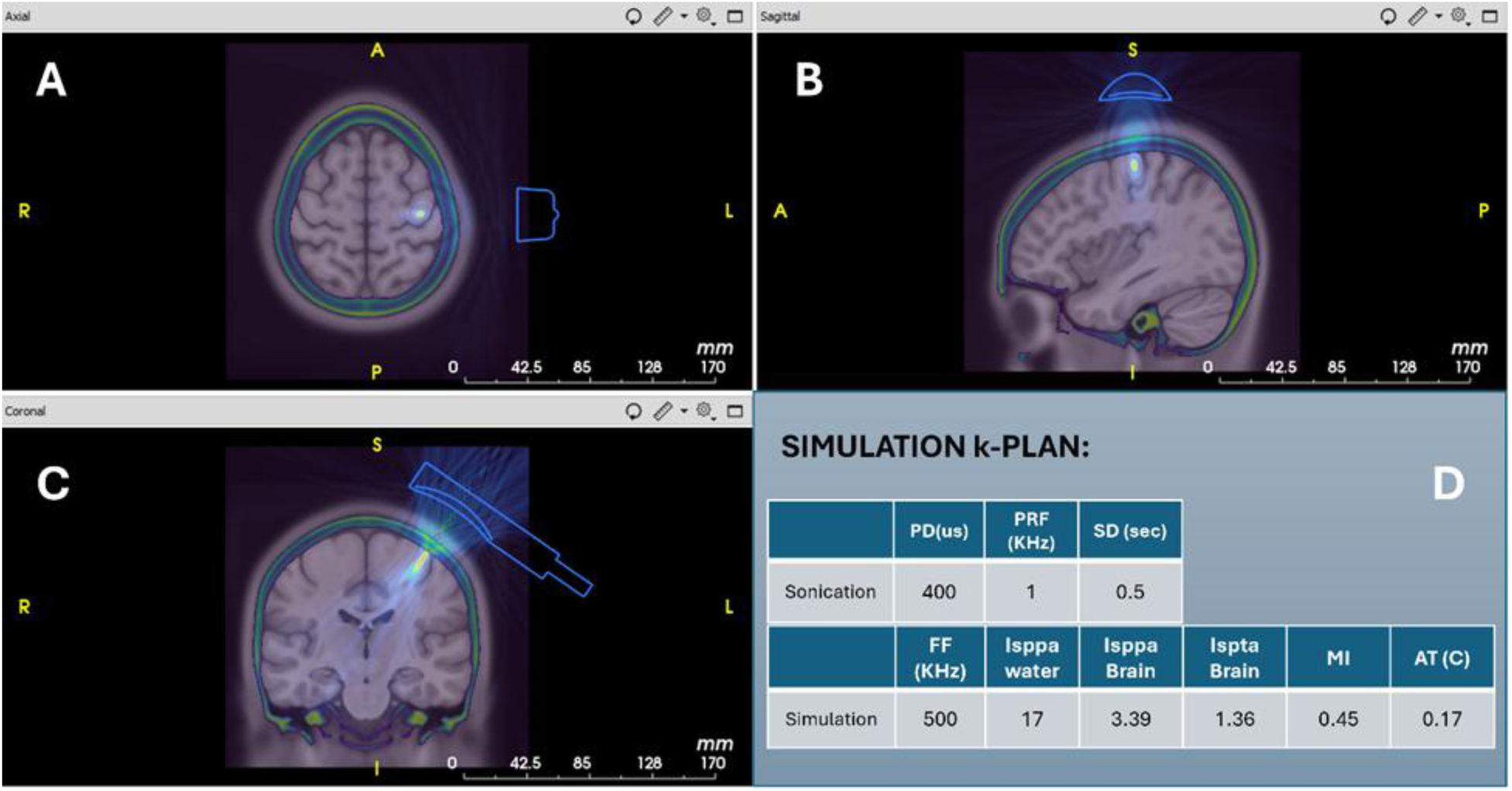
Low-intensity Transcranial Ultrasound Stimulation for Neuromodulation in Humans. (A, B, C) Example focal TUS targeting of a human motor cortex using k-plan software (BrainBox, Inc.). (D) TUS simulation software uses an input set of parameters (e.g., pulse duration, sonication duration, pulse repetition frequency, transducer fundamental frequency, intensity in water (ISSPA)) to simulate and calculate the estimated TUS intensity in the target brain region using the participant’s MRI and CT scans if available, or template human brain and CT scans. Simulation software will also generate the complete set of minimal parameters for reporting.

Ultrasound for clinical imaging or thermal ablation has a long history (Fry et al., 1951; Fry et al., 1950; Gavrilov, 1974). However, low-intensity ultrasound for neuromodulation remains a relatively nascent approach for non-invasive brain stimulation. Therefore, much remains to be understood about the mechanisms of TUS neuromodulation. Yet, considerable research progress has been made with TUS in humans and nonhuman animal models, narrowing the range of possible mechanistic hypotheses.

### Candidate mechanisms for TUS neuromodulation

Low intensity TUS in animal models has been shown to interact with neural tissue via mechanical effects. The sonication wave either directly changes the permeability of ion channels within neuronal membranes, such as voltage-gated sodium, calcium and potassium channels (e.g., K2P, TRP and Piezo1), or it temporary mechanically alters the cell membrane properties (Liao et al., 2021). Several mechanisms have been proposed, including changes in membrane turgidity and the dynamics of lipid microdomains (Anishkin et al., 2014; Babakhanian et al., 2018; Petersen et al., 2016; Suki et al., 2020; Tyler, 2011). TUS also affects the coupling between neurons and glial cells (Oh et al., 2019). The combination of TUS mechanical effects tends to increase action potentials being elicited by excitatory and inhibitory neurons (Tyler, 2011; Yoo et al., 2022). TUS has been shown to be capable of inducing muscle contraction and limb or tail flicking when rodent motor cortex is stimulated with low to moderate intensities (Kim et al., 2020; Lee et al., 2018; Tufail et al., 2010). Similar motor responses have yet to be observed and reported in human and non-human primates (Darmani et al., 2022).

At low intensities for neuromodulation, TUS can influence neural tissue without causing substantial damage, heating or adverse effects, as reported in human and non-human primates (Gaur et al., 2020; Spivak et al., 2021; Verhagen et al., 2019). However, care should be taken with continuous stimulation protocols where the continuity of stimulation (duty cycle; see Box 1) is high (Roumazeilles et al., 2021; Verhagen et al., 2019). Overall, TUS does not appear to cause significant heating or cavitation of brain tissue when the intensity remains low, based on Mechanical and Thermal Index values and recommendations of use (see Box 1 and 2). Temperature changes for low-intensity TUS are commonly <1°C (Baek et al., 2017; Yoo et al., 2011), and thermal effects could at least minimally alter cell membrane capacitance during ‘online’ TUS. However, thermal effects alone are unlikely to play a considerable role for longer-lasting ‘offline’ TUS effects (Ozenne et al., 2020; Verhagen et al., 2019). The mechanism of action for the longer-lasting offline effects is not yet well understood. Because the offline effects can last tens of minutes, or in some cases hours, after the sonication period (Bault et al., 2023; Pasquinelli et al., 2019), offline effects likely engage neuroplasticity mechanisms, such as modulation of AMPA and NMDA glutamatergic receptors and post-synaptic Ca^2+^ mediated changes to receptor properties. Interestingly, TUS effects on neuronal NMDA receptors appear to be indirect via, for instance, TUS modulation of astrocytes that can influence neuronal plasticity (Blackmore et al., 2023). TUS pulsed at a theta (4-8 Hz) frequency (theta-burst TUS; tb-TUS) is being studied for its capability to induce LTP-like plasticity (Darmani et al., 2025; Oghli et al., 2023; Samuel et al., 2023; Samuel et al., 2022; Zeng et al., 2022). We further consider tb-TUS as part of ‘offline’ stimulation protocols at the end of section (I) below. Of importance for future clinical applications, the repeated use of TUS sessions does not appear to negatively impact on the integrity of brain tissue as assessed by MRI (Munoz et al., 2022).

### Directionality of TUS neuromodulation

There is substantial interest in understanding the conditions under which TUS could be used to bias the directionality of neuromodulatory effects on the targeted brain area and its network or on behavior (Blackmore et al., 2023; Mihran et al., 1990; Tsui et al., 2005; Zhang et al., 2023). To describe the directionality of neural effects, we use the terms enhancement or suppression throughout, reserving the terms excitation and inhibition for references to reports where it was possible to directly record from identified excitatory and inhibitory neurons with animal models.

Recordings from identified excitatory and inhibitory neurons during TUS with animal models provide important mechanistic insights, because the neuronal recordings can be combined with causal manipulation, such as blocking specific ion channels. For instance, previous studies with murine models have reported that short Sonication Durations (SD, <1 sec) can lead to net neural excitation (attributed to more action potentials being elicited by TUS in excitatory neurons), whereas longer sonication durations (> 1 sec) can lead to net neural suppression (i.e., more strongly driving inhibitory neurons) (Mihran et al., 1990; Tsui et al., 2005). Other TUS studies have suggested that higher sonication Pulse Repetition Frequencies (PRF >100 Hz) can lead to net excitation (Manuel et al., 2020; Zhang et al., 2023). The caveat is that many nonhuman animal studies are conducted under anesthesia, which can alter the balance of excitatory-inhibitory neuronal activity. By comparison, although human TUS studies are often conducted without anesthesia, access to single units (neurons) is only possible with specialist FDA or ethical board approved electrodes for clinical monitoring in neurosurgery patients, with which it is difficult or not possible to identify the recorded cell types. Moreover, there is a paucity of direct neuronal recording studies in humans during TUS.

Researchers with other non-invasive brain stimulation approaches, including TMS, are working to overcome similar challenges in identifying the directionality of effects on neurons and neuronal networks (Fitzgerald et al., 2006). TMS researchers now regularly apply higher duty cycles to tip the directionality of TMS neuromodulatory effects on cortical areas towards net excitation (i.e., potentiation). By contrast, low-duty cycle TMS pulses are associated with net inhibition (i.e., de-potentiation or suppression) of muscle potentials or motor cortical responses (Solomon et al., 2024). Therefore, although the effects of TMS and TUS on neurons and neural systems differ, a certain level of correspondence between the two techniques may be found, even if it is not expected, in parameters such as duty cycle causing net excitation or suppression of neural network function.

Research into TUS mechanisms and effects is informing computational modeling, which allows the more systematic exploration of TUS stimulation parameters, difficult to achieve with any one empirical study alone. In a computational *Neuronal Intramembrane Cavitation Excitation* (NICE) model developed to study activation and suppression effects on modeled excitatory and inhibitory neuronal populations, TUS effects were simulated as intramembrane cavitation causing changes in ion channel conductivity (Plaksin et al., 2016). Although it remains questionable whether intramembrane cavitation is a key mechanism for TUS, the NICE model stimulation simulation approach can be viewed as general and it allowed the researchers to simulate a broad set of TUS parameters, including TUS intensity and the continuity of stimulation (duty cycle), on modelled neuronal responses.

Box 1 summarizes the common TUS parameters and their measuring units. Key parameters are the average acoustic intensity (intensity spatial peak pulse average, ISPPA), intensity spatial peak time average (ISPTA), sonication duration, duty cycle (DC), pulse repetition frequency (PRF), thermal index (TI) and mechanical index (MI). Box 2 shows guidelines on the ultrasound parameter limits that human low-intensity TUS studies typically follow.

The NICE model was initially evaluated with a more limited set of the previously available data from human and nonhuman animal studies, and the model at that time showed a high level of predictability, albeit with a very small sampling of studies available for comparison. Increases in intensity (ISPPA, Box 1) and duty cycle (DC, Box 1) were argued to be able to tip the balance from suppression to activation in the modeled populations of excitatory and inhibitory neurons (Plaksin et al., 2016). Several reviews have now conducted case-by-case and ad-hoc comparisons of TUS parameters with the NICE model predictions, with mixed support for the NICE model (Ai et al., 2018; Dell’Italia et al., 2022; Forster et al., 2023b; Zhang et al., 2023). In Box 3, we collate a set of net enhancement versus suppression hypotheses linked to TUS parameters that may be able to bias the directionality of neural effects. These hypotheses are generated by the animal research studies discussed in this section. Therefore, the hypotheses are not solely generated by the NICE model and are further considered in Section I on the meta-analysis and resource provided as part of this report.

The uncertainty about the extent to which TUS can be used to enhance or suppress neurobiological function limits its research potential (Chen et al., 1997; Fitzgerald et al., 2006). Rather than exploring the TUS parameter space, many researchers opt to emulate the TUS parameters of prior studies reporting specific positive findings, limiting the necessary exploration of the entire parameter space necessary for a nascent field and contributing to an empirical and reporting bias. We recognize the complexity of neural circuits and systems and the limitations in aiming to evaluate predictions with a relative paucity of data in humans. However, we also recognize that stepwise progress and evaluation are needed as signposts in this research endeavor, not unique to TUS or other brain perturbation approaches with longer histories of use, e.g., invasive deep-brain electrical stimulation, TMS, tACS, tDCS; see Fig. 1 (Derosiere et al., 2020; Klink et al., 2021). Therefore, since there are now many tens of human TUS studies (Figure 3), to us it seems the time is ripe for further research signposts and a database to be established as an open resource that can help to accommodate growth in the field. There are now a range of reported behavioral and neurobiological outcomes with human TUS, ranging from eliciting somatosensory sensations with TUS applied to the somatosensory cortex, the enhancement or suppression of the threshold for motor-evoked potentials (MEPs) with TUS applied to motor cortex (including in combination with TMS), the perception of visual phosphenes or modulation of visual motion perception from TUS applied to the visual cortex, and mood improvement induced by TUS to the prefrontal cortex (these and others are summarized in Tables 1-3). With the accumulation of these and other human low-intensity TUS data, a more extensive review and meta-analysis than previously possible can now be conducted. This could be a needed step towards the next key evaluation period when samples sizes increase.

**Figure 3.**
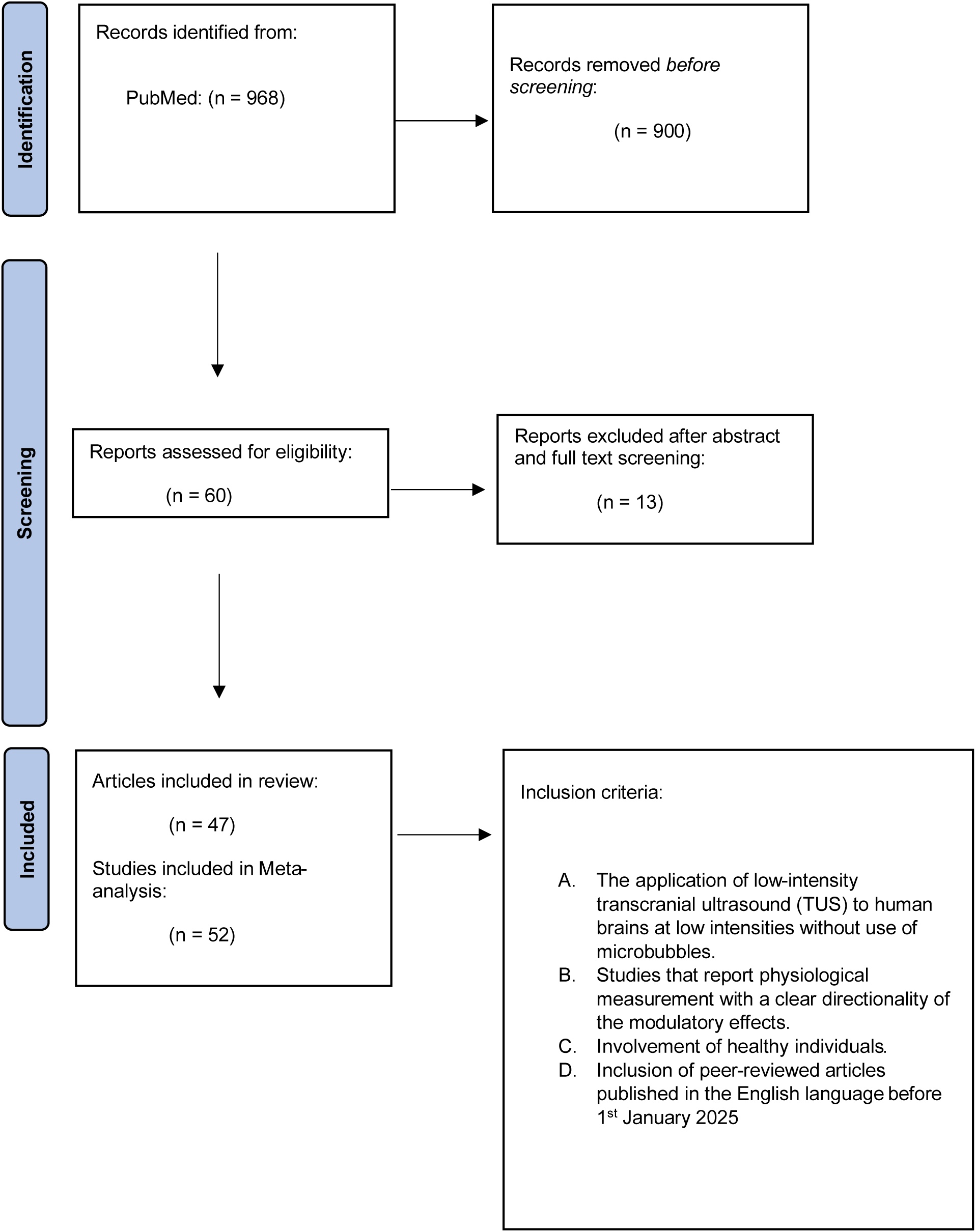
Meta-analysis selection and inclusion criteria using the PRISMA recommended approach. Selection and inclusion criteria for the meta-analysis with resulting sample sizes for the meta-analysis.

**Table 1.** Human ‘online’ TUS studies categorized by probable enhancement or suppression. Summarized are the TUS parameters reported in the human studies (inTUS_DATABASE_1-2025.csv) and their reported neurobiological effects categorized by likely excitatory or inhibitory effects. For independently confirmed effects, we cite the independent assessment source. *Abbreviations of Brain Targets:* Somatosensory Cortex (S1,S2); Primary Visual Cortex (V1); Primary Motor Cortex (M1); Ventro-Lateral Prefrontal Cortex (VLPFC); Ventral Capsule/Ventral Striatum (VC/VS); dorsal-Anterior Cingulate Cortex (dACC); Orbito Frontal Cortex (OFC); Thalamus-Ventral Posterolateral Nucleus (VPL); human Middle Temporal Area (hMT+); Nucleus Accumbens (NAcc); Medial Prefrontal Cortex (mPFCF); Anterior Insula (AI); Posterior Insula (PI); Entorhinal Cortex (ErC); *Abbreviations of Measurements:* Heartbeat Evoked Potential (HEP); Contact Heat Evoked Potential (CHEP); Heart Rate Variability (HRV); Transcranial Magnetic Stimulation (TMS); Short-Interval Intracortical Inhibition (SICI); Intracortical Facilitation (ICF); rs-fMRI (Resting State functional Magnetic Resonance Imaging); 3 Tesla fMRI (3T); Blood Oxygenation Level Dependent (BOLD); Electro Encephalography (EEG).

| ONLINE EFFECTS |  | PROBABLE ENHANCEMENT OF FUNCTION |  |  |  |  |  |  |  |  |
| --- | --- | --- | --- | --- | --- | --- | --- | --- | --- | --- |
| STUDY | TARGET | FF (kHz) | PRF (Hz) | DC (%) | SD (s) | PD (ms) | Isppa Water (W/cm<sup>2</sup>) | Isppa Brain (W/cm<sup>2</sup>) | Ispta Brain (W/cm<sup>2</sup>) | Reported Effect [independently characterized] |
| 1. (Lee et al., 2015) | Hand Region S1 | 250 | 500 | 50 | 0.30 | 1 | 3 | 0.70 | 0.35 | TUS enhanced somatosensory evoked potentials. [Wang et al. 2019] |
| 2. (Lee et al., 2016a) | S2 and S2+S1 | 210 | 500 | 50 | 0.50 | 1 | 35 | 8.80 | 4.40 | TUS elicited tactile sensations and enhanced somatosensory evoked potentials. [Wang et al. 2019; G. Darmani et al. 2022] |
| 3. (Lee et al., 2016b) | V1 | 270 | 500 | 50 | 0.30 | 1 | 16.60 | 11.60 | 5.80 | Simultaneous TUS and 3T fMRI resulted in increased BOLD signal in V1. With EEG: increased potentials (e.g., P100 and P200). Produced phosphene perception in some subjects. [Zhang et al. 2021] |
| 4. (Ai et al., 2016) Exp 1 (3T) | Sensorimotor | 500 | 1000 | 36 | 0.50 | 0.36 | 24 | 6 | 2.16 | TUS in sensorimotor cortex led to focal BOLD activation. [Yüksel et al.(2023)] |
| 5. (Ai et al., 2016) Exp 2 (7T) | Caudate | 860 | 1000 | 50 | 0.50 | 0.5 | 24 | 6 | 3 | TUS in caudate led to focal BOLD activation in subcortical areas. [Yüksel et al. (2023)] |
| 6. (Ai et al., 2018) | M1 (thumb) | 500 | 1000 | 36 | 0.50 | 0.36 | 16.95 | 4.23 | 1.52 | Increased fMRI BOLD signal in motor cortex (M1) thumb representation during a cued tapping task. [Wang et al. 2019; Biase et al. 2019] |
| 7. (Fine et al., 2020) | VLPFC | 500 | 1000 | 24 | 0.50 | 0.24 | 22.40 | 5.60 | 1.34 | TUS of right ventro-lateral PFC (VLPFC), enhanced frontal EEG potential (P300) for stop-trials in a stop signal task. (Enhanced inhibition action.) [Yüksel et al. (2023)] |
| 8. (Yu et al., 2020a) | M1 (leg area) | 500 | 3000 | 60 | 0.50 | 0.2 | 5.90 | 1.17 | 0.70 | Increased movement-related cortical activity when compared to sham, with the highest PRF used. [Yüksel et al. (2023)] |
| 9. (Liu et al., 2021) | S1 | 500 | 300 | 6 | 0.50 | 0.2 | 5.64 | 1.10 | 0.07 | TUS increased local excitability of S1 area EEG responses during sensory discrimination tasks. Early and late phase EEG potentials were enhanced. N300 potential amplitude was also enhanced. |
| 10. (Kuhn et al., 2023) | Entorhinal Cortex | 650 | 100 | 5 | 30 | 0.5 | 57.60 | 14.40 | 0.72 | Increased fMRI activity in entorhinal cortex and functional connectivity. |
| 11. (Zhang et al., 2023)--> Excitatory protocol | M1 (hand area) | 500 | 2000 | 40 | 0.50 | 0.2 | 2.46 | 0.62 | 0.25 | Excitatory protocol, combining TUS + TMS of motor cortex increased M1 excitability and decreased intracortical inhibition. |
| 12. (Chou et al., 2025)<br>Protocol A | VC/VS | 650 | 10 | 5 | 30 | 5 | 57.6 | 14.4 | 0.720 | Augmented putamen activation during reward task. Increased rs-fMRI functional connectivity with ACC and OFC. |
| **AVERAGE** |  | **490.8** | **909.2** | **34.3** | **5.4** | **0.9** | **22.6** | **1.8** | **6.2** |  |

**Table 1. Human ‘online’ TUS studies categorized by probable enhancement or suppression.**
| ONLINE EFFECTS PROBABLE SUPPRESSION OF FUNCTION |  |  |  |  |  |  |  |  |  |  |
| --- | --- | --- | --- | --- | --- | --- | --- | --- | --- | --- |
| STUDY | TARGET | FF (kHz) | PRF (Hz) | DC (%) | SD (s) | PD (ms) | Isppa Water (W/cm<sup>2</sup>) | Isppa Brain (W/cm<sup>2</sup>) | Ispta Brain (W/cm<sup>2</sup>) | Reported Effect [independently characterized] |
| 13. (Legon et al., 2014) | S1 | 500 | 1000 | 36 | 0.50 | 0.36 | 23.9 | 6 | 2.2 | TUS to S1 attenuates somatosensory evoked potentials. Improved tactile discrimination task performance. |
| 14. (Mueller et al., 2014) | S1 | 500 | 1000 | 36 | 0.50 | 0.36 | 23.9 | 6 | 2.2 | Disrupted intrinsic brain activity and phases for beta, not for gamma. [Zhang et al. 2021] |
| 15. (Legon et al., 2018a) | Thalamus VPL | 500 | 1000 | 36 | 0.50 | 0.36 | 14.5 | 7.03 | 2.53 | Attenuation of somatosensory evoked component. Attenuation of alpha and beta power. [Zhang et al. 2021] |
| 16. (Legon et al., 2018b) | M1 | 500 | 1000 | 36 | 0.50 | 0.36 | 17.12 | 4.28 | 1.54 | Attenuation of motor evoked potential during single-pulse TMS. Attenuation of intracortical facilitation (ICF), unaffected SICI. |
| 17. (Fomenko et al., 2020) | M1 | 500 | 1000 | 10 | 0.50 | 0.1 | 9.26 | 2.32 | 0.232 | TMS motor evoked potential suppression in a dose-dependent manner and increase in short interval intracortical inhibition. [Zhang et al. 2021; Yüksel et al. (2023)] |
| 18. (Cain et al., 2021b)--> Mode 1 | Globus Pallidus | 650 | 100 | 5 | 30 | 0.5 | 57.6 | 14.40 | 0.72 | TUS decreased fMRI BOLD signal and within the brain network observed. |
| 19. (Xia et al., 2021) | M1 | 500 | 1000 | 30 | 0.50 | 0.3 | 9.26 | 2.32 | 0.69 | Inhibition of ipsilateral M1 potential after TUS. |
| 20. (Butler et al., 2022) | hMT+ | 500 | 1000 | 50 | 0.30 | 0.5 | 5.76 | 1.44 | 0.72 | TUS to visual area hMT+ produced attenuation of visual motion EEG evoked potentials. |
| 21. (Kuhn et al., 2023) | Amygdala | 650 | 10 | 5 | 30 | 5 | 57.6 | 14.40 | 0.72 | Decreased functional inter-connectivity with amygdala. |
| 22. (Zhang et al., 2023) --> Inhibitory TUS | M1 (hand area) | 500 | 50 | 2 | 0.50 | 0.4 | 2.46 | 0.62 | 0.01 | Inhibitory protocol: TUS + TMS --> Increased short and long interval intracortical inhibition with reduced intracortical facilitation. |
| 23. (Peng et al., 2024) | NAC | 650 | 10 | 5 | 30 | 5 | 57.52 | 14.38 | 0.719 | TUS on left NAC inhibited activity of anterior and posterior regions of bilateral NAC, compared to sham. Increased functional connectivity between NAC and mPFC after TUS. |
| 24. (Strohman, Isaac, et al., 2024) | AI, PI | 500 | 1000 | 36 | 1 | 0.36 | 4.2 | 1.05 | 0.378 | TUS to PI produced reduction of EEG HEP relative to sham and relative to effects of TUS to dACC, in the 280ms post R-peak, 650ms post R-peak time windows, respectively. |
| 25. (Legon et al., 2024) | AI, PI | 500 | 1000 | 36 | 1 | 0.36 | 3.5 | 0.88 | 0.31 | TUS to AI and PI reduced amplitude of CHEP.<br>TUS to AI increased HRV.<br>TUS to PI reduced pain perception. |
| 26. (Chou et al., 2025)<br>Protocol ErC | ErC | 650 | 100 | 5 | 30 | 0.5 | 57.6 | 14.4 | 0.720 | Decreased putamen activation during reward task.<br>Decreased ErC perfusion. |
| **AVERAGE** |  | **542.9** | **662.1** | **23.4** | **9.0** | **1.0** | **24.6** | **1.0** | **6.4** |  |

Our key objectives with this report are twofold. In Part I, we evaluate the initial set of collated net neural enhancement versus suppression hypotheses (Box 3), and we conduct an initial meta-analysis of the available healthy human participant low-intensity TUS reports. We conclude Part I by establishing an open Iowa-Newcastle (inTUS) resource, including a database that can grow with TUS research community input and an analysis script that can (re)analyze the data. The inTUS resource is offered to encourage TUS researchers to report outcomes on a broader and more systematic exploration of the TUS stimulation parameter space. The reporting approach that we use is as recommended by the International Transcranial Ultrasonic Stimulation Safety and Standards (ITRUSST) consortium, which has proposed standards to enable comparison and reproducibility across studies (Martin et al., 2024), also see https://itrusst.com.

In Part II of this report, we selectively summarize the current state of the literature on human TUS applications for perturbing the brain and developing treatments for neurological and psychiatric disorders. This literature review is not intended to be a comprehensive review of the TUS literature on clinical applications. More comprehensive reviews of this literature can be found elsewhere (Matt et al., 2024; Pellow et al., 2024). Instead, this selective summary of TUS in specific clinical disorders aims to illustrate how hypotheses on the directionality of TUS effects need to be patient and brain area/system specific. Therefore, at the conclusion of each of the Part II sections, we motivate testable hypotheses on how TUS could be employed to enhance or suppress function for that patient-specific application. We encourage the reader to continue to develop hypotheses, not limited to the examples provided here.

### Part I. Net enhancement and suppression hypotheses and meta-analysis

In this section, we, 1) consider the rationale for the hypotheses regarding the directionality of TUS effects (Box 3); 2) overview the approach for the meta-analysis; and, 3) discuss the initial analyses and results obtained with the meta-analysis of the database, including the limitations of the current approach. We conclude by establishing the database and analysis script as an inTUS community resource that can grow. The inTUS resource includes the inTUS_DATABASE tables and an R markdown script to recreate the analyses as the database grows in R Studio. To make the resource more interactive and user friendly, even without relying on R or R Studio, the user can study the results via an HTML summary of the results and figures and browse TUS parameter interactions via an online app. The inTUS resource aims to support the more systematic exploration and reporting of the TUS parameter space and outcomes, as recommended by the ITRUSST consortium (Martin et al., 2024). We hope that the TUS community will be able to contribute to growing the database for further hypothesis development, empirical testing and modeling.

### Net enhancement and suppression hypotheses

The hypotheses summarized in Box 3 are based on TUS parameters that have been highlighted in the TUS literature to be capable of biasing TUS effects on a given brain region towards neural enhancement or suppression. These hypotheses were generated both from the NICE model (Plaksin et al., 2016; Plaksin et al., 2014) and preclinical studies with animal models. The preclinical studies of TUS effects have reported conditions under which TUS could potentially bias the directionality of effects towards greater excitation or inhibition. The NICE model hypothesized that two key parameters associated with net activation or suppression (using the authors’ terminology) are sonication intensity in the target brain area (ISSPA in brain) and the continuity of stimulation (duty cycle, DC). The NICE model predictions are shown in Figure 4 with a light blue line defining the border between neural enhancement (higher DC and intensity) and suppression (lower DC and intensity). Other parameters of interest are ISPTA, which mathematically multiplies ISPPA by the continuity of sonication (DC), and the length of the sonication pulse (Sonication Duration, SD). Shorter SDs (<500 ms) have been reported to elicit more action potentials from excitatory neurons, and longer SDs (>500 ms) tend to elicit suppression via greater excitation of *inhibitory* neurons (Mihran et al., 1990; Tsui et al., 2005). Other studies have suggested that pulse-repetition frequency (PRF), the frequency with which the ultrasound pulse is turned on/off, can bias towards greater net excitation at higher PRFs (Kim et al., 2023; Yu et al., 2020). These hypotheses are summarized in Box 3.

**Figure 4.**
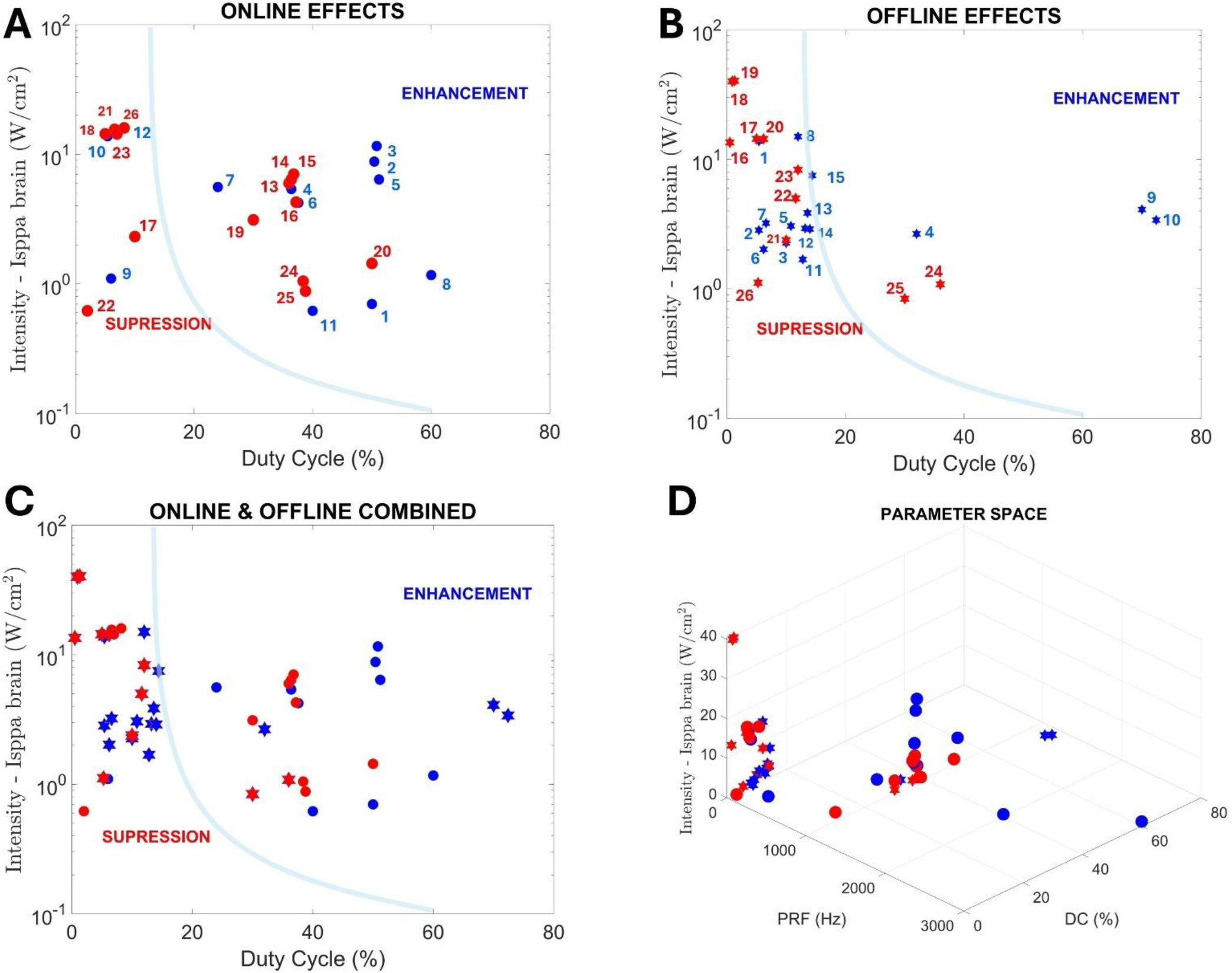
Online and offline TUS study parameters and outcomes. (A) Online effects, based on the studies in Table 1, are shown segregated by probable enhancement versus suppression. Light blue line shows the NICE model boundary between suppression and enhancement. Blue circles are human studies reporting probable enhancement, red circles probable suppression. Index numbers correspond to the studies numbered in Table 1. (B) Same format and analysis approach showing the “offline” effects studies in Table 2 with stars. Numbers correspond to the studies numbered in Table 2. (C) Combined figure with online (Table 1) and offline (Table 2) studies. Same symbol and color use as in A-B. (D) Additional hypothesized parameters, such as pulse repetition frequency (PRF), can be plotted from the inTUS_DATABASE in multi-dimensional spaces as shown and with the inTUS resource app.

**Table 2.**
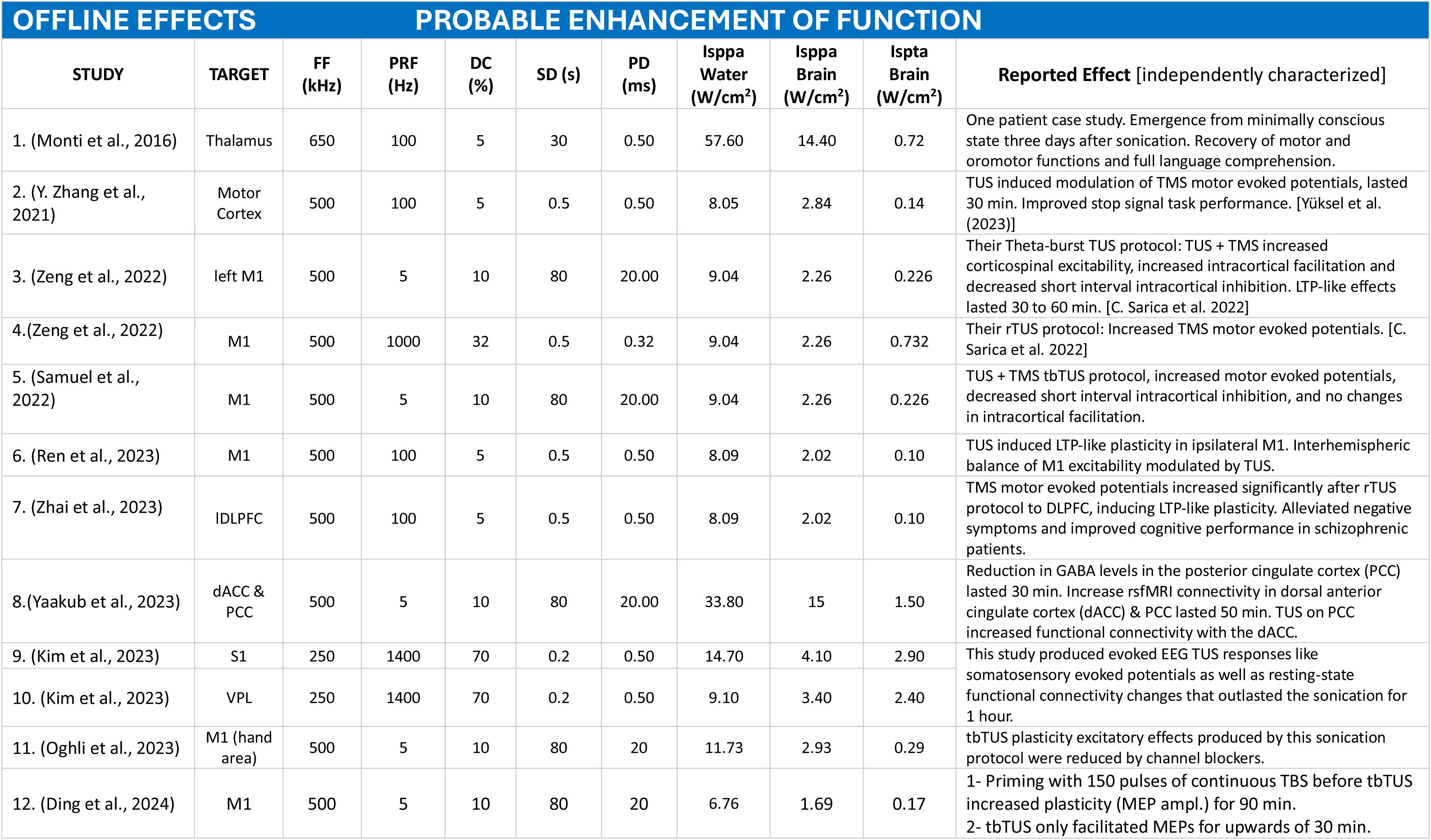
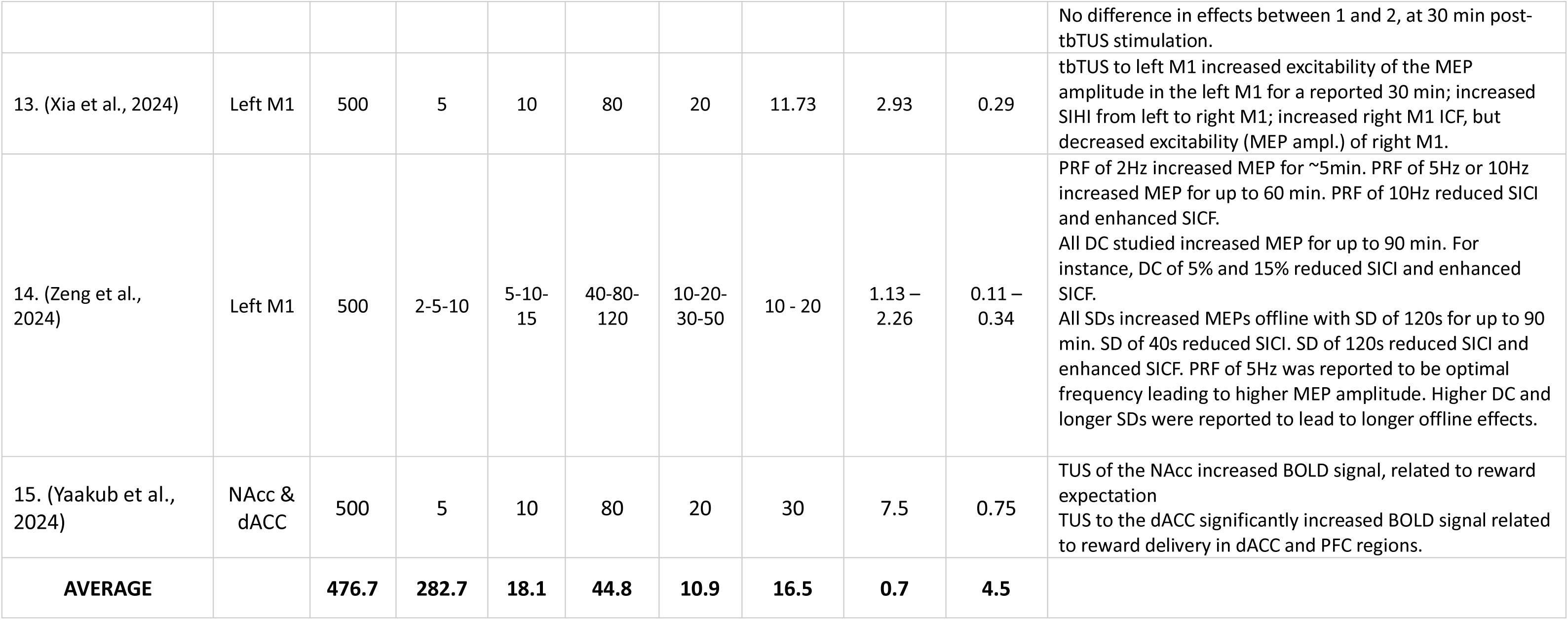
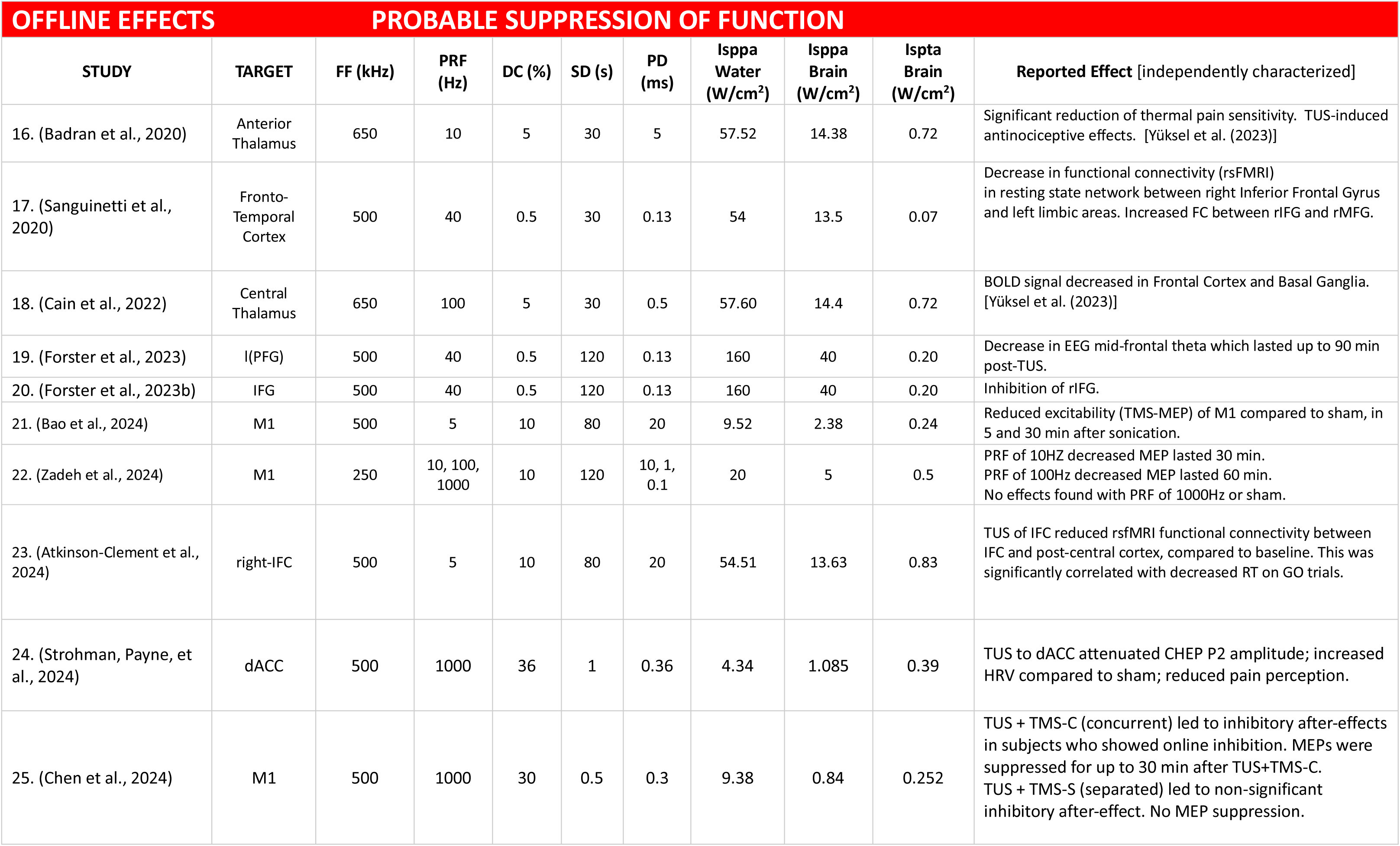
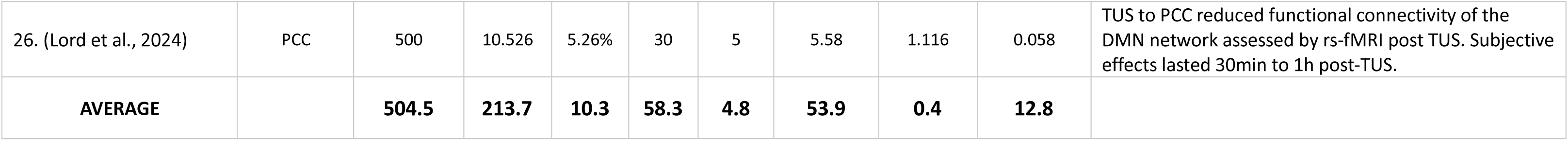
Human ‘offline’ TUS studies categorized by probable enhancement or suppression. Summarized are the TUS parameters used in human studies (inTUS_DATABASE_1-2025.csv) aiming to induce longer lasting ‘offline’ effects. Format as in Table 1. *Abbreviations of Brain Targets:* Dorso-Lateral Prefrontal Cortex (DLPFC); Primary Motor Cortex (M1); dorsal-Anterior Cingulate Cortex (dACC); Posterior Cingulate Cortex (PCC); Somatosensory Cortex (S1); Ventral Posterolateral Nucleus (VPL); Nucleus Accumbens (NAcc); Inferior-Prefrontal Gyrus (IPFC); Inferior Frontal Gyrus (IFG); right-Inferior Frontal Gyrus (rIFG); right-Medial Frontal Gyrus (rMFG). *Abbreviations of Measurements:* Transcranial Magnetic Stimulation (TMS); Short-Interval Interhemispheric Inhibition (SIHI); Intracortical Facilitation (ICF); Short-Interval Intracortical Inhibition (SICI); Short-Interval Intracortical Facilitation (SICF); Long-Term Potentiation (LTP); repetitive-Transcranial Ultrasound (rTUS); theta-burst TUS (tbTUS); resting-state functional Magnetic Resonance Imaging (rs-fMRI); Blood Oxygenating Level Dependent (BOLD); Electroencephalography (EEG); Motor Evoked Potential (MEP); Contact Heat Evoked Potential (CHEP); Reaction Time (RT); Heart Rate Variability (HRV); Default Mode Network (DMN).

### Segregating ‘online’ and longer lasting ‘offline’ effects

Studies of online or offline effects tend to use different TUS parameters (compare Tables 1 and 2). Offline effects of TUS stimulation are those induced for longer time periods after the sonication period (seconds or minutes). Because many human studies aim to keep the intensity of offline stimulation within FDA and/or ITRUSST guidelines (Box 2), a different parameter range for online and offline studies may be implemented by TUS researchers. Namely, for offline studies, DC and ISPTA values are often kept low (Figure 4B). For this reason, we summarize the online and offline studies in separate tables (Tables 1-2) and include this distinction as a factor in the database meta-analyses.

### Meta-analysis inclusion criteria and analysis approach

The meta-analysis approach follows the Preferred Reporting Items for Systematic reviews and Meta-Analyses (PRISMA) guidelines (Figure 3). We searched PubMed/MEDLINE for studies published through 1^st^ January 2025 by employing the combination of the following keywords: ‘human’, ‘ultrasound’, ‘focused’, ‘low-intensity’, ‘stimulation’, ‘transcranial’, ‘neuromodulation’, ‘TUS’, ‘FUN’, ‘LIFUS’ ‘clinical’, ‘treatment’ (see Figure 3 for the papers in the initial sample). Four authors (HC, NS, CP and MZ) searched for and curated the included studies to ensure that the survey was as comprehensive as possible. We searched only for articles in English. Study inclusion criteria for the meta-analysis focused on human studies involving low-intensity TUS for brain stimulation or neuromodulation. The included studies encompassed only healthy human participants to avoid including clinical studies of patients with brain pathology at this stage of the database development, although we intend to include patient clinical studies in future versions of the database. We excluded studies with moderate-intensity or high-intensity ultrasound used for, respectively, BBB perturbation or thermal ablation, outside of the scope of the current version of the database. Given the currently small size of the database, we expected the analyses not to be robust to missing data. For this reason, we had to exclude 4 diagnostic ultrasound studies (Gibson et al., 2018; Guerra et al., 2021; Hameroff et al., 2013; Schimek et al., 2020), which did not report the parameters we needed for the meta-analysis. After applying the inclusion and exclusion criteria, 32 studies were included in the first inTUS_DATABASE_6-2024, and 47 studies were included in the next key expansion of the database: inTUS_DATABASE_1-2025. Further key database expansions, analyses and results will be reported in the inTUS resource location noted below. For the meta-analysis, we only included studies that either reported a basic set of TUS stimulation parameters or those sufficient for estimating the required parameters necessary for the meta-analysis.

The reported studies’ TUS parameters and reported effects were used to populate the data tables (Table 1 for online studies; Table 2 for offline studies). Most parameters required for the analysis could be found in the reported studies or calculated from the parameters given. If a given study conducted multiple complete experiments, the sample sizes reflect the overall number of experiments rather than the number of studies/papers, and if separate experiments tested different values of a given parameter with the same result, the experiment eliciting the strongest effect was input into the database for meta-analysis. A number of experiments (n = 14 out of 37 in the inTUS_DATABASE_6-2024, and an additional, n = 13 out of the 15 additional experiments added in the inTUS_DATABASE_1-2025), although reporting ISPPA values in water, did not report ISSPA values in the brain, required for our analyses, which is a recognized problem for this field (Martin et al., 2024). For these studies, we applied an accepted approximation of ISPPA values, because the sonic wave passes through and loses much of its energy at the skull, whereby typically 70-75% of the intensity is lost (Fomenko et al., 2020; Lee et al., 2015; Mueller et al., 2014; Oghli et al., 2023; Xia et al., 2024; Zeng et al., 2024). We compared these approximated values entered into the database to simulations using k-plan software (Jaros et al., 2020; Treeby & Cox, 2010), targeting the same regions as in the reported experiments with the reported ISPPA in water values. Comparing the k-plan simulation values to the estimated values showed a low margin of error of ∼5% between the two sets of values in the comparisons. Therefore, we used the approximated values for studies not reporting ISPPA in the brain.

Probable net neural enhancement versus suppression was characterized as follows. Note that our use of the terms enhancement and suppression refers exclusively to the increase or decrease of neural activity, respectively, as measured by, neurophysiological methods (EEG-ERPs, BOLD fMRI, etc.) and do not imply equivalent changes in behavioral responses. Although many studies have reported behavioral influences, these alone were not sufficient for us to determine the probable directionality of underlying neural TUS effects. Although many TUS studies report using a sham control (e.g., no TUS or an inverted TUS transducer that directs the sonic energy away from the head), it is difficult to rule out other sources for placebo effects in the behavioral reports. We thereby focused on the studies’ reporting of neurobiological effects and characterized these effects as probable net neural enhancement versus neural suppression, using the following approach. For net enhancement, we followed the prior approach established by the TMS field, whereby EEG-evoked responses that are magnified in the target area as a function of TUS application could be characterized as probable enhancement (see Tables 1-2). We included positive fMRI BOLD effects resulting from TUS as a probable enhancement. For suppression, we also followed the prior approach established by the TMS field, whereby EEG evoked responses that were reduced indicate likely suppression of neural activity. Wherever possible, we relied on independently characterized directionality of effects, and we cite the original sources that conducted the characterization in Tables 1 and 2. As an example of a study categorized as ‘neural suppression’ of function, Legon and colleagues (Legon et al., 2014) examined TUS targeting the primary somatosensory cortex (S1) in healthy human subjects with EEG. The authors reported that TUS significantly attenuated somatosensory evoked potentials. The effects were specific to the targeted region, because the changes were abolished when the acoustic beam was focused away from S1. As another example, Liu and colleagues applied TUS to S1 of participants performing a sensory discrimination task, and they reported augmented somatosensory spatiotemporal EEG responses, which are interpreted as increased local excitability or ‘neural enhancement’ by our terminology (Liu et al., 2021).

The resulting database tables were submitted to an exploratory univariate logistic regression model tested with R Studio. The R markdown script used to generate the results from the data tables is shared as part of the inTUS resource below. The sample sizes in the initial inTUS_DATABASE_06-2024 were 37 experimental observations of each of the TUS parameters in the database. The sample sizes in the subsequent inTUS_DATABASE_01-2025 were 52 experimental observations. The first statistical model tested the Enhancement versus Suppression Effect separately for ISPPA, Duty Cycle, PRF and SD from the hypotheses with the Mode of stimulation ‘online’ or ‘offline’ as a covariate (e.g., logit ∼ OfflineOnline + Isppa, etc.). A single model with all factors and all interactions would have been preferred, but with these sample sizes is not sufficiently powered to provide meaningful results. This will be revisited in the future when sample sizes increase through the database expansion via the inTUS resource. For the second key database expansion period (inTUS_DATABASE_01-2025), we also conducted an initial multivariate pattern analysis using a Random Forest Walk analysis implemented in python (Scikit-learn: Machine Learning in Python) (Pedregosa et al., 2011).

### Human TUS meta-analysis results

The meta-analysis used the data in Tables 1 and 2 from the inTUS_DATABASE_1-2025.csv. The tables summarize the range of TUS parameters of interest for the studies reporting probable enhancement or suppression of TUS effects. The rationale for characterization of the directionality of TUS effects, independently evaluated wherever possible, is summarized in Tables 1-2 in the rightmost column. The database is further separated by studies aiming to elicit online (Table 1) or offline effects (Table 2). Further key evaluation periods of the inTUS database, as it expands, may not be reported with updates to this paper, but will continue to be reported online in the inTUS resource. The statistical models, statistical output variables and resulting figures are available and can be recreated in the inTUS resource for any of the datasets available via the R markdown script.

### Results from initial *inTUS_DATABASE_6-2024*

This section reports the results from the first database analysis (inTUS_DATABASE_6-2024.csv), with a sample size of n = 37 experiments (from 33 studies). The statistical model for DC was significantly associated with the Enhancement versus Suppression, our effect of interest (*p* = 0.046). Sonication duration (Thurman et al.) was also significantly associated with the effect of interest (SD: *p* = 0.040). Consistent with the hypotheses in Box 3, higher DC values were significantly associated with Enhancement (n = 21; mean DC during Enhancement = 29.952, standard deviation (STD) = 23.252), lower values with Suppression (n = 16; mean DC during Suppression = 16.719; SD = 17.159). Also consistent with the hypotheses in Box 3, lower SD brain values were significantly associated with Enhancement (n = 21; mean SD = 14.6190, STD = 28.736) and higher values with suppression (n = 16, mean SD = 29.612, STD = 41.369). A statistical trend was present for the Online and Offline covariate factor (*p* = 0.061), as might be expected given the different parameters that are often used for online and offline studies (Figure 4a-b).

There was no significant effect for ISSPA brain (*p* = 0.256), and the other parameters of interest were also not significantly associated with our effect of interest in this dataset: pulse repetition frequency (PRF: *p* = 0.324) or ISPTA (*p* = 0.787). Therefore, in this initial dataset evaluation, only DC and SD were significantly associated with Enhancement versus Suppression in the hypothesized direction (Box 3). No multivariate pattern analyses were attempted with this dataset.

### Results from inTUS_DATABASE_1-2025

This section reports the results from the *second* key database expansion meta-analysis (inTUS_DATABASE_1-2025.csv), with a sample size of n = 52 experiments (from 47 studies) as shown in Tables 1-2. The exploratory univariate statistical model for DC (*p* = 0.077) and for ISSPA brain (*p* = 0.097) showed a statistical trend in their association with the Enhancement versus Suppression effect of interest. Consistent with the hypotheses in Box 3, higher DC values trended in association with Enhancement (n = 27; mean DC during Enhancement = 25.333, standard deviation (SD) = 22.231), lower values with Suppression (n = 25; mean DC during Suppression = 17.630; SD = 16.052). Inconsistent with the hypotheses in Box 3, *lower* ISPPA brain values trended in association with Enhancement (n = 27; mean ISPPA brain during Enhancement = 5.276 W/cm^2^, standard deviation (SD) = 4.696), higher values with Suppression (n = 25; mean ISPPA brain during Suppression = 9.221; SD = 10.718).

The Online and Offline study covariate factor was not significantly different in any of the statistical models (*p* > 0.1). Rerunning the statistical models separately on the Online and Offline effects did not result in any of these parameters showing a significant association or a statistical trend, consistent with the effects not being statistically distinguishable in the current database (1-2025). This might be either because the sample sizes are not sufficiently powered for statistical effects to be observed or the TUS community might be converging somewhat in the TUS parameters used for online or offline stimulation (Fig. 4). The remaining parameters of interest were not significantly associated with the Enhancement versus Suppression effect of interest: PRF (*p* = 0.324) and sonication duration (SD: p = 0.326).

For this database, we conducted an initial multivariate pattern analysis using a Random Forest Walk procedure (53 observations, 4 variables: PRF, DC, SD, ISPPA brain, from Tables 1 and 2). The mean accuracy of effect classification (200 times 5-fold cross-validation) with all these variables was 0.55 (SD = 0.14), compared to analysis with a shuffled target variable (TUS effect directionality: Enhancement vs Suppression; mean 0.48 and SD = 0.15), see Figure 6. Although the accuracy of the database predicting the effect of interest is numerically above chance, the SD values overlap the original and shuffled data, and the permutation importance of the parameter values (Figure 6, right panel), are low (between 0.04 and –0.06). These observations suggest that the database sample size is too small to provide a meaningful result with this multivariate analysis at this stage. The classification accuracy was similar when all variables from Tables 1-2 were included in the analysis (8 variables: FF, PRF, DC, SD, PD, ISPPA_water, ISPPA_brain, ISPTA_brain; classification accuracy 0.57, SD = 0.14; shuffled accuracy 0.50, SD = 0.14).

### Impressions from the *inTUS_DATABASE_1-2025* results

Given the still limited sample size of human TUS studies to date, we interpret these meta-analysis results with caution. A key observation is that the NICE model is not as strongly predictive as initially thought (Dell’Italia et al., 2022; Zhang et al., 2023). Nonetheless, in the exploratory univariate analyses, DC and ISPPA are statistical trends for an association with the categorical Enhancement versus Suppression variable in the database. The DC effect is in the hypothesized direction. However, the ISPPA_brain effect is not (see Box 3; compare Figure 5 for DC and ISSPA_brain). Namely, consistent with the hypotheses in Box 3, higher DC values trended in association with Enhancement and lower values with Suppression. Inconsistent with the hypotheses in Box 3, lower ISPPA brain values trended in association with Enhancement and higher values with Suppression (see Figure 5). The multivariate pattern analyses with the four parameters of interest classified at an accuracy level close to chance and the factor importance values were too small to be meaningfully interpreted (Fig. 6). In relation to the initial dataset results (inTUS_DATABASE_6-2024), the DC factor which was significant in the first is now a trend in the second analysis. The ISPPA_brain factor was not significant previously with the univariate analyses, but is a trend with the second dataset evaluated (inTUS_DATABASE_1-2025). Therefore, the effects are still labile and may change, warranting caution in over-interpretation at this stage.

**Figure 5.**
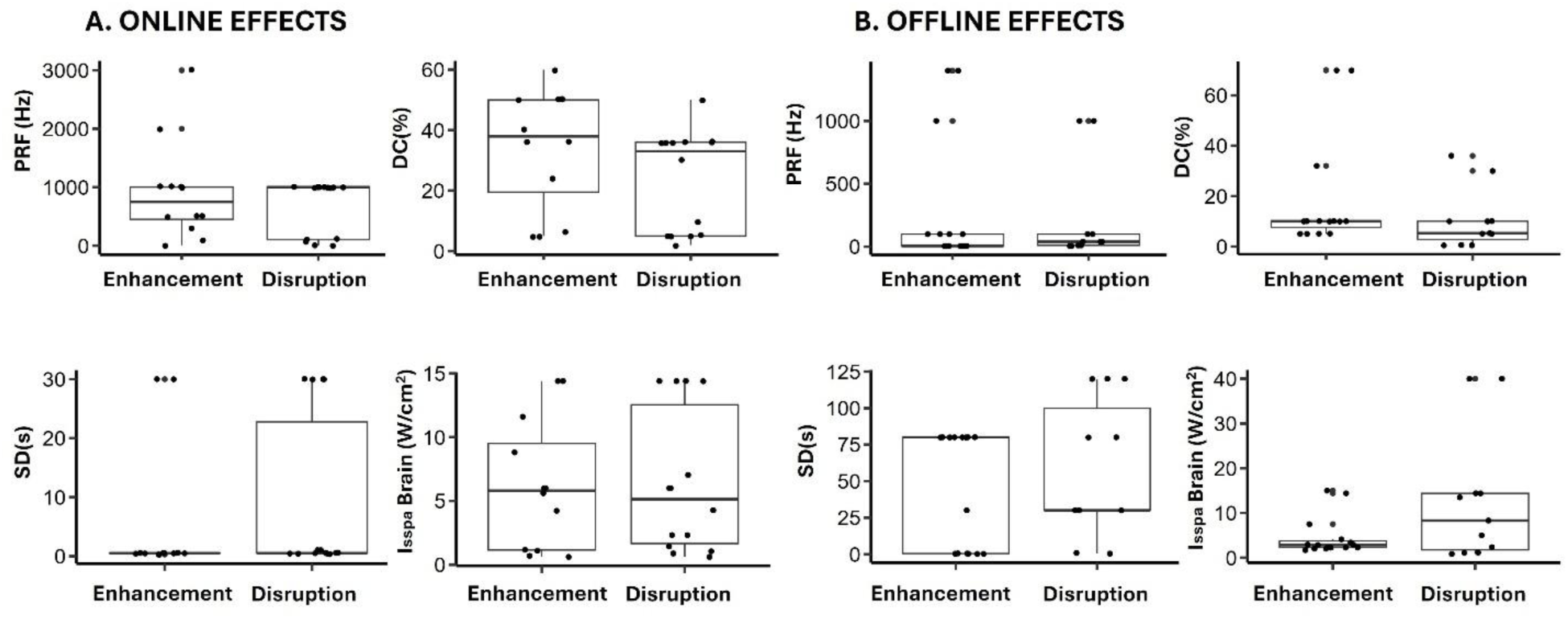
Box plots of meta-analysis results for the TUS parameters of interest. Shown are boxplots for TUS parameters of interest segregated by probable enhancement or suppression (data from Tables 1 and 2). Plots show TUS parameters: Pulse Repetition Frequency (PRF), Duty Cycle (DC), Sonication Duration (Thurman et al.) and ISPPA Brain. These are shown separately for Online effects (A) and Offline effects (B).

**Figure 6.**
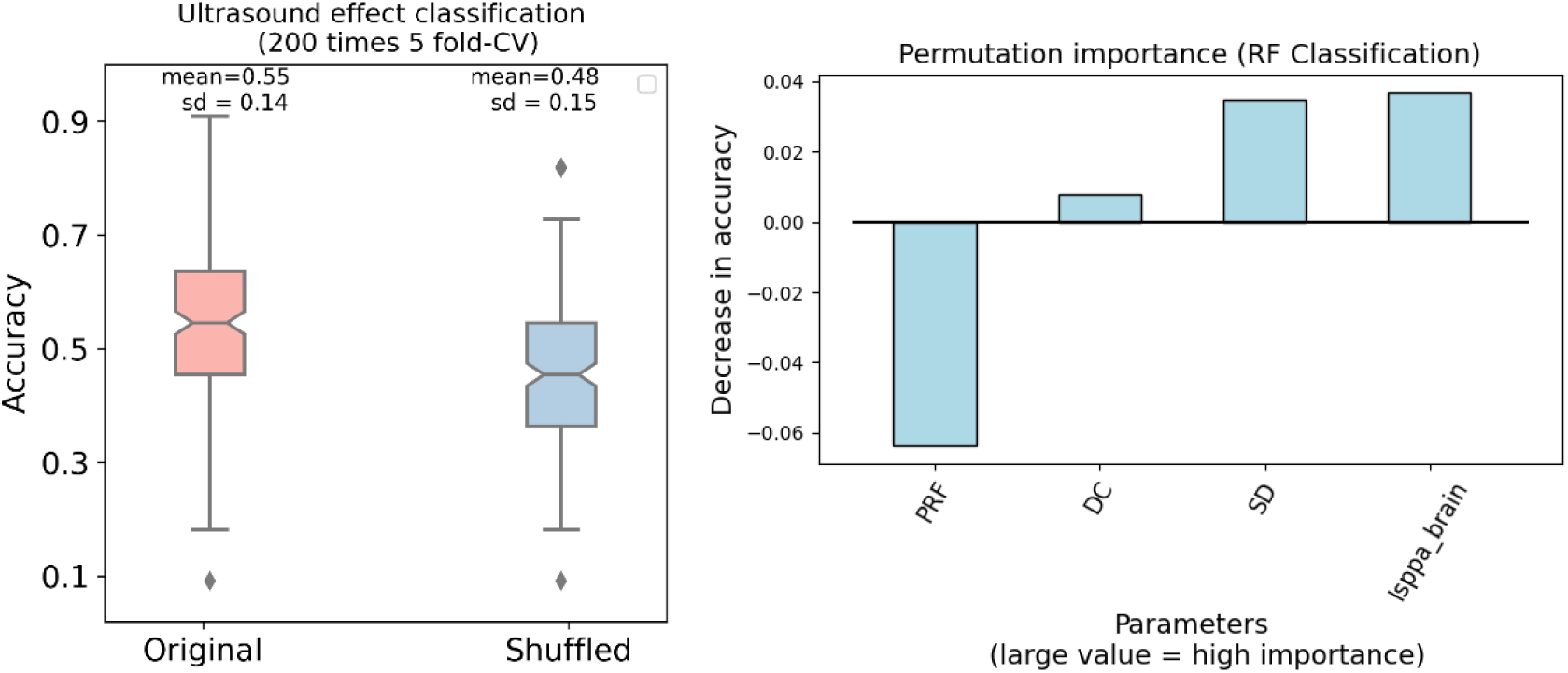
Random Forest Walk multivariate pattern analysis results. Results of the Random Forest analysis with the 4 parameters of interest in the second key database inTUS_DATABASE_1-2025 compared to a shuffled parameter dataset (left). Left panel shows the average accuracy (mean = 0.55; SD = 0.14) is not far from the shuffled dataset (mean = 0.48; SD = 0.15), suggesting that the dataset size may not yet be robust for multivariate analyses. The right panel shows the ordered decrease in accuracy when the variable is permuted (permutation importance: higher values carry more importance). We observe that the importance values are low (between –0.06 and 0.04; right panel), warranting caution in interpretation at this stage of the inTUS database

### Theta Burst TUS for offline neuromodulation

Theta-burst TUS (tb-TUS) is being studied for its capability to induce cortical LTP-like plasticity for an extended period of time (Darmani et al., 2025; Oghli et al., 2023; Samuel et al., 2023; Samuel et al., 2022; Zeng et al., 2022). Theta-burst TUS protocols are identified in Table 2. We observe that tb-TUS protocols tend to consist of a more continuous stimulation approach than online protocols, such as 80s trains of 20ms sonication pulses spaced over 200ms (i.e., pulsed at a 4-8 Hz theta frequency). However, independent replication of these findings remains limited. For example, Bao *et al*., found reduced motor cortex excitability – measured as decreased TMS-MEP amplitude in M1 -- that lasted up to 30 minutes post-sonication (Bao et al., 2024). Whereas Fong *et al*. reported no significant effects between tbTUS and sham conditions in M1 excitability (Fong et al., 2024). These studies, although growing, are still too few to consider separately, and as such are currently included as ‘offline’ studies in the database. Of note, some groups are seeing enhancement and others disruption with their tb-TUS protocols (Table 2). As the number of tb-TUS studies grows in the database, it will be important to evaluate tb-TUS outcomes and compare these to other TUS stimulation protocols.

### Meta-analysis limitations

A key limitation of this meta-analysis is the relatively small sample size. Multi-variate analyses of these data could have more predictability with greater sample sizes to provide deeper insights into the interplay of TUS parameters and effects. Moreover, some effects with certain parameters of interest have statistically changed between the initial (6-2024) and second (1-2025) key dataset evaluation periods reported here. Therefore, as noted above, the effects remain labile but may stabilize with future dataset expansion and evaluation periods. Currently it is unclear which if any TUS parameters or sets of parameters together will be good predictors for controlling the directionality of TUS effects. Another limitation is the inherent selection bias of retrospective studies, whereby researchers may limit their exploration of the TUS parameter space based on studies with positive findings. Researchers may also be limiting their brain targeting options, and there may be a positive finding reporting bias. Moreover, we and others have noted that not all TUS researchers share the full set of key parameters necessary for secondary hypothesis testing, even though the ITRUSST consortium has recommended that TUS studies report a basic set of parameters like those included in the inTUS database (Martin et al., 2024). For instance, we had to simulate the derated ISPTA values in the brain, which although comparable to our k-plan simulation values warrant caution when interpreting results. Therefore, the results from the 6-2024 and 1-2025 databases analyses need to be considered as purely explorative. For this reason, we have not evaluated the predictability of the models, which the statistical tests at this stage suggest is low.

We will continue to periodically report updates to these results in the inTUS resource, and the database and results can be re-analyzed at any time using the R markdown script in the freely available software R Studio. We hope that the inTUS resource will encourage more systematic exploration and reporting of the TUS parameter space, complemented with computational modeling to fill gaps in the empirical research. Other caveats worth mentioning are that the TUS community is relatively small, and many researchers work with several TUS research groups around the world. It is important to be able to identify independent replication studies. In the online version of the table, which can expand as needed, we have included additional columns that could be used for future analyses, including a column to identify independent replication or contradiction studies. Another important facet is that the current database has focused on healthy participants. Patient studies and those in animal models (e.g., rodents and nonhuman primates) would be important to include in future iterations of the database, and we welcome input on growing the database via the inTUS Qualtrics survey. Moreover, in the online data table we have added a column to identify cortical versus subcortical sites that were stimulated. Currently most TUS studies in the database have focused on cortical stimulation. These additional columns could be used for further analysis and hypothesis testing as the database grows.

### How to use and contribute to the *inTUS Resource*

To help to address the noted limitations to the current database, we establish the inTUS Resource as an open database and a set of analysis tools. The inTUS Resource can be found in the Open Science Framework link here: https://osf.io/arqp8/, in the folder associated with this paper (Caffaratti_et_al_inTUS_Resource folder). This repository contains the data tables at each of the key evaluation periods (currently 6-2024 and 1-2025) and the R markdown script to regenerate the statistical tests, results and figures using the freely available R or R Studio software (Posit). The resource also has a link to a Qualtrics form that researchers can use to submit parameters and outcomes as part of their preprint or published work. A more detailed description of the inTUS resource contents, how they are curated and how to contribute to the resource can be found as a READ_ME file in the resource documents page, a copy of which we have appended at the end of this report. Please cite this paper if you use or rely on the inTUS Resource.

As noted here, not all studies are reporting the range of ITRUSST recommended TUS stimulation parameters. We encourage TUS researchers to contribute more accurate values to the database from their studies and to more systematically report the more complete set of values as recommended by the ITRUSST consortium (Martin et al., 2024). This will allow the data to be mined more completely, which may further support or refute these or other hypotheses. We will retain the historic datasets on the resource pages as a point of reference. We welcome input via the online form on improving the criteria for assessing neurobiological or behavioral effects, which will benefit the entire TUS community and is a key objective of the ITRUSST consortium https://itrusst.com/. The original OSF link noted here will point to the location of the repository even if it moves location.

### Meta-analysis summary

Given the sample size limitations, these retrospective meta-analysis results are tentative, with the possibility that the results may stabilize, strengthen or change. The combination of the meta-analysis and resource are made openly available as an inTUS Resource to further support and encourage the TUS research community to more systematically report TUS parameters and study outcomes. Furthermore, we encourage the TUS research community to explore the full TUS parameter space whenever safe and possible to conduct in human and nonhuman animals or rely on computational modeling for exploration of the extreme reaches of the TUS parameter space. We fully anticipate that this effort will dovetail with a need for further computational modeling and empirical study in human and nonhuman animals. TUS effects could also in the future benefit from being modeled across the cortical depth, in interaction with other brain areas (Thorpe et al., 2024) or insights on the impact on cell and molecular properties of the neural network.

### Part II. Selective TUS clinical applications review and TUS directionality hypotheses

Compared to pharmaceutical drugs that can affect many parts of the brain and body, TUS allows the stimulation of specific targets within the brain with relatively high spatial precision. Here, we selectively review potential applications for low-intensity TUS that are currently investigational. We also selectively consider moderate-intensity applications for Blood Brain Barrier (BBB) perturbation and high-intensity TUS for clinical thermal ablation. While the primary goal of BBB opening is to regionally increase the permeability of the BBB to enhance the efficacy of brain drug delivery, BBB opening alone could induce neuromodulatory effects (Chu et al., 2015). Our purpose with the below selective review is not to provide another comprehensive review of the TUS clinical application literature (Matt et al., 2024; Pellow et al., 2024). Instead, our purpose here is to illustrate how hypotheses on the directionality of TUS effects could be developed, which we do not intend to be limited to the examples provided here. The reader will notice that for some applications there are no or few TUS studies, whereas for other applications there are more. Nonetheless, for each example application in the low-intensity TUS section where directionality of effects on neuromodulation could be useful to harness, we have aimed to motivate at the end of each section example testable hypotheses that could be further developed. We hope the reader finds this exercise useful, because TUS clinical applications and the directionality of effects need to be specifically tailored for the pathologies and brain regions/network being targeted.

### Ia. Low-intensity TUS applications

#### Motor and somatosensory system mapping

Intraoperative clinical motor and somatosensory cortical mapping is important for planning neurosurgical treatment. TMS over the motor cortex is regularly used to induce muscle contractions and limb movements. The effect of TMS on the amplitude of muscle-evoked potentials is an accepted measure of motor cortical enhancement (increased motor-cortical evoked EEG potentials) or suppression (decreased MEPs) (Fitzgerald et al., 2006). In preclinical research, low- or moderate-intensity TUS focused on motor cortex in rodents can induce muscle contractions (King et al., 2014; Tufail et al., 2010; Yoo et al., 2011; Younan et al., 2013), including limb, tail, whisker or eye muscle contraction (King et al., 2014). TUS in humans targeting the motor cortex can either enhance or suppress MEPs (Table 1) (Gibson et al., 2018; Lee et al., 2016a; Legon, Bansal, et al., 2018; Samuel et al., 2022; Xia et al., 2021; Zeng et al., 2022; Y. Zhang et al., 2021). Stimulation of the primary motor cortex with TUS has been found to decrease reaction times in a stimulus-response task, which is interpreted as enhanced motor performance (Fomenko et al., 2020; Legon, Bansal, et al., 2018; Zhang et al., 2022; Y. Zhang et al., 2021).

For mapping human somatosensory cortex, TUS has been reported to either enhance or suppress somatosensory evoked potentials (SEPs) recorded with EEG, and TUS can elicit a range of somatosensory perceptions, such as tactile sensations in the hand contralateral to the stimulated somatosensory cortex (Lee et al., 2015; Legon et al., 2014). Legon *et al*. demonstrated impaired performance in a tactile spatial discrimination task from TUS stimulation of the ventro-posterior lateral nucleus of the thalamus (Legon, Ai, et al., 2018). This effect was related to the disruption of the corresponding SEP component (Legon, Ai, et al., 2018). *<u>Hypothesis</u>:* For clinical motor or somatosensory cortical mapping, TUS could induce movement behavior or somatosensory percepts by stimulating motor/somatosensory sites. Alternatively, TUS could be used to suppress ongoing motor functions (hand squeeze, arm drop).

#### Speech and language mapping

Intra-operative brain mapping using electrical stimulation is used by neurosurgeons to identify brain areas crucial for speech and language (Benzagmout et al., 2007; E. F. Chang et al., 2015; Duffau, 2010; Mandonnet et al., 2017; Mathias et al., 2016). The gold-standard approach identifies speech and language areas using electrical stimulation to elicit speech arrest, naming or other language difficulties (Duffau, 2010; Mathias et al., 2016). However, because of the limited time in the operating room for patient brain mapping, there is considerable interest in developing pre-operative non-invasive brain stimulation approaches for speech and language brain mapping. For instance, TMS, when used with MRI-based neuro-navigation to target neocortical speech and language regions, can lead to speech arrest or anomia, which generally corresponds to the locale of intra-operative mapping using electrical stimulation (Tarapore et al., 2013). Furthermore, TMS is often integrated with adjunctive methodologies such as electroencephalography (EEG), functional magnetic resonance imaging (fMRI), or diffusion tensor imaging (DTI) to bolster the precision and specificity of brain-behavioral mapping. To date there do not appear to be TUS studies focused on speech and language mapping, defining a clear research need. *<u>Hypothesis</u>:* For speech and language mapping, TUS could temporarily suppress brain areas important for speech production and language function, analogous to the current use of electrical stimulation for intra-operative mapping of neocortical areas involved in these processes.

#### Mood disorders

TUS has been explored as a possible treatment for psychiatric mood disorders. In a study from 2013, in humans with chronic pain, TUS administered to the posterior frontal cortex contralateral to the source of pain elicited a significant mood enhancement after 40 minutes (Hameroff et al., 2013). Sanguinetti *et al*. reported that TUS targeting the right ventrolateral prefrontal cortex led to reports of improved mood in healthy individuals after TUS (Sanguinetti et al., 2020). In a double-blind pilot study, Reznik and colleagues applied TUS to the right fronto-temporal cortex of depressed patients, resulting in mood improvement (Reznik et al., 2020; Shimokawa et al., 2022). Forster *et al*. used TUS to indirectly manipulate cingulate cortex activity in a learned helplessness task, demonstrating the potential to affect the response to acute stressors that can induce symptoms of depression (Forster et al., 2023). Further pre-clinical and clinical trial studies would be necessary to evaluate TUS efficacy in alleviating mood disorder symptoms. *<u>Hypothesis</u>:* TUS could be used to target highly interconnected brain network hubs associated with depression risk or resilience (Trapp et al., 2023) to potentially modulate mood bidirectionally.

#### Schizophrenia

Early pilot results for patients suffering from psychosis are now available. In a double-blind, randomized, sham-controlled study, 15 sessions of TUS over the left dorsolateral prefrontal cortex (DLPFC) could alleviate negative symptoms in schizophrenia patients and enhance cognitive performance in a continuous performance test (Zhai et al., 2023). TUS was well tolerated with patients in the active group, which did not report more adverse effects than patients in the sham group. The use of TUS seems particularly promising due to the involvement of deep-brain structures, such as the thalamus in this condition (Mukherjee & Halassa, 2024). *<u>Hypothesis</u>:* For TUS application to schizophrenia, TUS could suppress the function of target areas, reducing the positive or negative symptoms associated with schizophrenia.

#### Disorders of Consciousness

Low-intensity TUS has shown the capability to hasten the recovery of behavioral responsiveness in patients with disorders of consciousness (Lee et al., 2016a). Monti and colleagues documented a case where low-intensity TUS aimed at the thalamus was associated with the emergence from a minimally conscious state in patients experiencing disorders of consciousness following severe brain injury (Monti et al., 2016). *<u>Hypothesis</u>:* For this clinical application, TUS could enhance the function of thalamic nuclei and interconnectivity with other brain areas, such as the centro-median-perifascicular nuclei of the thalamus and the mesencephalic reticular formation (Chudy et al., 2023). However, it is worth noting that a more permanent approach, such as electrical deep brain stimulation (DBS), may be required in some patients or a combination of TUS ‘mapping’ followed by DBS.

#### Alzheimer’s disease

Cognitive decline associated with dementia would benefit from approaches that can enhance cognitive function. In a study with 11 Alzheimer’s disease (AD) patients using transcranial pulse stimulation (TPS; typically shorter pulses of low-intensity ultrasound stimulation over a longer period of time) targeting the hippocampus, the authors reported that 63% of patients improved on one or more cognitive assessment (Nicodemus et al., 2019). In another study involving 35 AD patients, TPS was applied to the dorsolateral prefrontal cortex (Beisteiner et al., 2020). The patients’ neuropsychological scores significantly improved after TPS, and these improvements were reported to have persisted for up to three months. However, TPS studies remain highly limited and would require further study and comparison to effects with other TUS protocols. *<u>Hypothesis</u>:* TUS for AD applications could enhance the function of target regions to enhance or stabilize neural system inter-connectivity and cognitive function.

#### Parkinson’s disease

In a study by Nicodemus *et al*. 11 patients took part in research with TUS for Parkinson’s Disease (PD) targeting the substantia nigra. In this study, 87% of the patients had either stable or improved fine motor scores and 88% had stable or improved gross motor scores (Nicodemus et al., 2019). Samuel *et al*. used a technique called accelerated theta-burst TUS targeting the primary motor cortex in 10 PD patients, studying its impact on neurophysiological and clinical outcomes (Samuel et al., 2023). Their patients received both active and sham TUS conditions, and the authors measured TMS-elicited motor-evoked potentials (MEPs) before and after treatment. The study found a significant increase in TMS induced MEP amplitudes following TUS but not sham treatment. *<u>Hypothesis</u>:* For applications related to PD, TUS of the subthalamic nucleus could suppress its function in a lasting way if targeting the motor segment of the nucleus can be conducted with precision to avoid limbic or sensory parts of the nucleus.

#### Epilepsy

TUS application to an epileptogenic site has the potential to modulate seizure frequency. To evaluate these possibilities, Lee *et al*. applied low-intensity TUS to individuals with drug-refractory epilepsy undergoing intracranial electrode monitoring with stereo-electroencephalography (SEEG) (Lee et al., 2022). Two of the six patients studied showed a *decrease* in seizure occurrences, while one experienced an *increase*. The TUS effects reported were at electrode contacts near to the subsequent neurosurgical treatment site for epilepsy. Across all frequency bands in the local field potential recorded from the SEEG electrodes, there was a notable decrease in spectral power for all six patients following TUS. However, there was no clear relationship between these immediate effects on interictal epileptiform discharges and alterations in seizure frequency (Lee et al., 2022). Another study introduced a device for delivering pulsed low-intensity TUS to the hippocampus in humans, with no reported adverse events after multiple sessions (Brinker et al., 2020). A study by Bubrick *et al*. described the application of serial TUS in patients with mesial temporal lobe epilepsy. TUS was delivered in 6 sessions over 3 weeks. No adverse events or side effects were reported. Early results were promising, with significant seizure reduction in 5 out of 6 patients reported for up to 6 months after TUS application (Bubrick et al., 2024). *<u>Hypothesis</u>:* For epilepsy treatment, TUS could reduce the probability of seizures. However, for clinical mapping of epileptogenic sites, clinical mapping during epilepsy monitoring procedures will depend on TUS inducing epileptiform activity.

<u>Sudden unexpected death in epilepsy</u>. Sudden unexpected death in epilepsy (SUDEP) refers to the sudden unexpected death of a person with epilepsy that cannot be explained by trauma or status epilepticus. On post-mortem examination, no structural or toxicological cause of death can be ascertained. SUDEP is one of the leading causes of premature deaths in epilepsy, accounting for more than a 20-fold increase in the risk of sudden death in epileptic patients compared with the general population (Ficker et al., 1998; Kløvgaard et al., 2022). Among all neurological conditions, it ranks second after stroke in terms of years of potential life lost (Thurman et al., 2014). Rare cases of SUDEP of patients in epilepsy monitoring units have shown that cessation of breathing (apnea) following seizures precedes terminal asystole and death (Bateman et al., 2008; Nashef & Brown, 1996; Ryvlin et al., 2013). Animal models (Johnston et al., 1995) confirm a primary role of respiratory dysfunction in SUDEP. In the human patient work by Dlouhy and colleagues (Dlouhy et al., 2015; Harmata et al., 2023; Rhone et al., 2020), it was shown that when a circumscribed site in the amygdala, referred to as the Amygdala Inhibition of Respiration (AIR) site, is affected either by the spread of seizure or by electrical stimulation, apnea occurs without the patient feeling any air hunger or alarm (Lacuey et al., 2017; Nobis et al., 2019). In a subsequent study by Harmata et al., 2023, electrical stimulation or stimulation evoked seizure within a focal region of the AIR site evoked apnea that persisted well beyond the end of stimulation or seizure. Because this site in the amygdala caused persistent inhibition of respiration, the authors referred to this site as the pAIR site. The AIR site, proposed to be a brain region that mediates seizure-induced inhibition of breathing, which can persist for minutes and may lead to SUDEP.

Localization and characterization of the AIR site and pAIR site have so far been done using electrical stimulation in patients who have electrodes implanted for potential surgical remediation of epilepsy. This puts a severe constraint; only a limited population of individuals with epilepsy who are candidates for electrode implantation have contributed to the characterization of these sites. Extension to a larger population of epileptic patients without amygdala implantation and non-epileptic patient controls require the use of non-invasive methods. Given the deep subcortical location of the AIR and pAIR sites, approaches such as TMS are less suitable for this purpose. Because TUS has the capability to target deep areas with higher spatial resolution, it can be used to target not only the AIR sites in the amygdala, but also the respiratory network underlying SUDEP. Although there is a pressing research need for TUS research into controlling SUDEP, we could not find TUS studies on SUDEP in the literature. *<u>Hypothesis</u>:* TUS could suppress amygdala function to prevent apnea. However, rather than controlling SUDEP risk during epileptic seizures, if TUS cannot induce lasting effects or be implemented continuously, its utility may be better suited to identify people at high risk for SUDEP based on seizure-associated apnea. Namely, TUS could be used to stimulate the AIR site to confirm its location for subsequent neurosurgical ablation, for example, to reduce or eliminate epilepsy patient SUDEP risk.

#### Stroke and neuroprotection in brain injury

Low-intensity TUS has been studied for its potential neuroprotective benefits following brain injury and stroke by boosting neurotrophic factors as well as angio-neurogenesis (increased brain derived neurotrophic factor (BDNF), increased vascular endothelial growth factor (VEGF) and reduced apoptosis) (Bretsztajn & Gedroyc, 2018; Ichijo et al., 2021; Schellinger et al., 2015; Su et al., 2017; Tufail et al., 2010; Yang et al., 2015).Chen *et al*. **s**timulated mice with TUS before inducing cerebral ischemia and reported , reporting improved neurological function and decreased neuronal cell apoptosis (Chen et al., 2018).

In a randomized controlled trial, Wang *et al*. investigated the effects of TUS combined with cognitive rehabilitation on post-stroke cognitive impairment (Wang et al., 2022). The group receiving both TUS and cognitive rehabilitation exhibited improvement in a range of cognitive measures compared to the group that received cognitive rehabilitation alone. (Wu et al., 2019)This type of work suggests that TUS could be a potential preventive therapy for recurrent stroke, presuming it can have longer lasting effects after stimulation or be delivered (semi)continuously as needed. (Hanawa et al., 2014; Ichijo et al., 2021; Imashiro et al., 2021)TUS has also been explored as a non-invasive thrombectomy tool to enhance thrombolysis with tissue plasminogen activator in acute stroke (Schellinger et al., 2015). For stroke thrombectomy, TUS would act to break up thrombocytes with or without a tissue plasminogen activator. An earlier study, by Liu *et al*., indicated that administering TUS soon after a stroke could yield neuroprotective effects (Liu et al., 2019). Thus, there has been interest in evaluating whether initiating TUS promptly post-stroke could effectively enhance cerebral blood flow, revive local circulation, save the ischemic penumbra, and minimize brain tissue harm. However, TUS appears to have also led to a higher incidence of cerebral hemorrhage in patients concurrently treated with intravenous tissue-type Plasminogen Activator, tPA (Daffertshofer et al., 2005; Katsanos et al., 2020; Schellinger et al., 2015) .*<u>Hypothesis</u>:* TUS applications for stroke recurrence could enhance neuroplasticity and vascular perfusion, or they can be used to improve thrombolysis after acute stroke.

#### Hypertension and cardiovascular system effects

As a promising noninvasive therapy for drug-refractory hypertensive patients, Li and colleagues demonstrated the antihypertensive effects and protective impact on organ damage by using low-intensity TUS stimulation in spontaneously hypertensive rats (Li et al., 2023). The experiment involved daily 20-minute TUS stimulation sessions targeting the ventrolateral periaqueductal gray in the rats for two months. Their results showed a significant reduction in systolic blood pressure, reversal of left ventricular hypertrophy, and improved heart and kidney function. The sustained antihypertensive effect may be attributed to the activation of antihypertensive neural pathways and the inhibition of the renin-angiotensin system. Ji and colleagues explored the feasibility of using low-intensity TUS to modulate blood pressure in rabbits (Ji et al., 2020). The study used a TUS system to stimulate the left vagus nerve in rabbits while recording blood pressure in the right common carotid artery. Different TUS intensities were tested, showing a decrease in systolic and diastolic blood pressure, mean arterial pressure and heart rate (Ji et al., 2020). The higher the TUS intensity, the more significant the blood pressure reduction. These pre-clinical studies in animal models highlight the possibility of non-drug management of hypertension using TUS, opening avenues for treating clinical hypertension non-invasively. *<u>Hypothesis</u>:* TUS for hypertension application could suppress sympathetic nodes (e.g., rostral ventro-lateral medulla) or enhance parasympathetic nodes (e.g., medial prefrontal cortex) in the central autonomic network (Macefield & Henderson, 2020; Shoemaker, 2022).

#### Ib. Moderate intensity TUS applications

For moderate intensity TUS application, we do not generate directionality of TUS effects hypotheses. This section is only intended to provide a brief introduction to moderate intensity TUS applications, mainly to distinguish them from low-intensity TUS neuromodulation.

#### Enhancing pharmacological- and immuno-therapy through the blood-brain barrier

A significant challenge in drug- or immune-therapy is the limited effectiveness of drugs and vectors that do not easily traverse the blood-brain barrier (BBB) (Hynynen et al., 2006; Mehta et al., 2021), an issue that has been explored in the context of using antibodies to amyloid β to treat Alzheimer’s disease. TUS has the ability to temporarily open the BBB, facilitating the entry of vectors into the brain from the blood stream. Systemic injection of microbubbles when combined with TUS temporarily opens the BBB, with BBB integrity restored within 4–6 hours (Hynynen et al., 2006; Mehta et al., 2021). Lipsman and colleagues conducted a phase I safety trial, using TUS to safely and reversibly open the BBB in five patients diagnosed with early to moderate AD (Lipsman et al., 2018). They achieved predictable BBB opening at approximately 50% of the power at which cavitation was observed during a test using the NeuroBlate system. Right after the ultrasound treatment, a distinct rectangular-shaped enhancement was visible in the targeted brain region on T1-weighted gadolinium MR images. This enhancement was resolved within 24 hours after the procedure, suggestive of successful closure of the BBB. The moderate intensity TUS did not lead to any significant clinical or radiographic adverse events, nor a noticeable decline in cognitive scores at the three-month follow-up when compared to baseline. Importantly, no serious adverse events, such as hemorrhages, swelling, or neurological deficits were reported either on the day of the procedure or during the follow-up study period. Rezai *et al*. employed TUS to breach the BBB in a study involving six AD patients (Rezai et al., 2020). Post-treatment contrast-enhanced MRI scans displayed rapid and significant enhancement in the hippocampus, which subsequently resolved. Throughout the several TUS treatments, no adverse effects were observed, and there was no cognitive or neurological functional decline. In a study by Jeong *et al*. involving four AD patients, moderate-intensity TUS of the hippocampus did not exhibit evidence of actively opening the BBB, as observed in T1 dynamic contrast-enhanced MRI (Jeong et al., 2021; Jeong et al., 2022). However, the authors found that the regional cerebral metabolic rate of glucose (rCMRglu) in the superior frontal gyrus and middle cingulate gyrus significantly increased following TUS treatment. The patients also demonstrated mild improvement in measures of cognitive function after TUS, including memory function. Although BBB opening could lead to neuromodulatory effects, its effects at the network level are distinct from those achieved with TUS for neuromodulation (Liu et al., 2023).

#### Ic. High-intensity ultrasound for thermal ablation

For high intensity TUS application, we do not generate directionality of TUS effects hypotheses. This section is only intended to provide a brief introduction to high intensity TUS applications and to distinguish these from low-intensity TUS.

#### Parkinson’s disease

Moser *et al*. introduced high-intensity MR-guided TUS for thermal ablation as a potential treatment option for Parkinson’s disease, employing it to target and ablate the connections between the thalamus and globus pallidus (Moser et al., 2013). Their approach improved the patients’ Unified Parkinson’s Disease Rating Scale (UPDRS) score by 57%. This potential therapeutic benefit of high-intensity ultrasound for PD was further underscored by Magara *et al*. in 2014, who used MR-guided TUS to thermally ablate the unilateral pallidothalamic tract in PD patients, resulting in significant improvement in the UPDRS score three months post-surgery (Magara et al., 2014).

#### Essential tremor

TUS at high intensities that cause tissue ablation has FDA-approved application for essential tremor following large, randomized clinical trials (Choi & Kim, 2019; Krishna et al., 2018). Precision thermal ablation of subthalamic nuclei is increasingly considered as an alternative to deep brain stimulation for select patients (Rohani & Fasano, 2017). MR-guided TUS is being employed in treating essential tremor (ET) with the thalamic ventral intermediate (VIM) nucleus as the primary target (Abe et al., 2020; W. S. Chang et al., 2015; Elias et al., 2013; Elias et al., 2016; Lipsman et al., 2013; Meng et al., 2018). This thalamotomy technique has demonstrated therapeutic benefits for essential tremor patients and has received FDA approval for unilateral treatment (Elias et al., 2016). The reported side effects of thermal ablation with high-intensity TUS include early symptoms of dizziness, nausea/vomiting, headache, skull overheating, flushing, and late symptoms such as ataxia and paresthesia (Abe et al., 2020; W. S. Chang et al., 2015; Elias et al., 2013; Elias et al., 2016; Lipsman et al., 2013; Meng et al., 2018).

#### Epilepsy

In a recent case report, MR-guided high-intensity TUS was found to be effective in a patient with medically intractable epilepsy, resulting in 12 months of seizure freedom (Abe et al., 2020). For a more extensive review of TUS for thermal ablation in epilepsy patients see (Cornelssen et al., 2023).

## BOXES

**Box 1.**
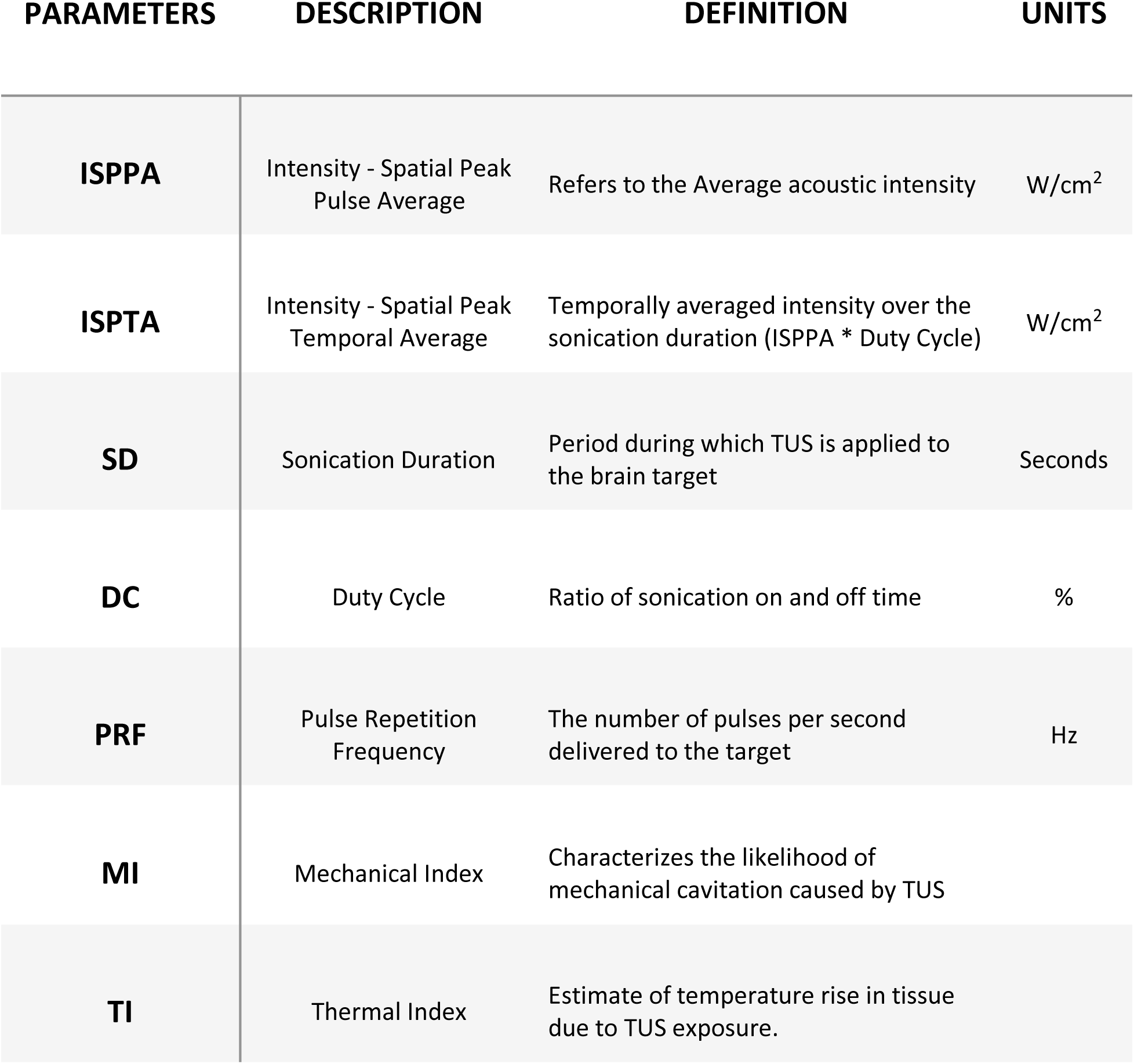
Transcranial focused ultrasound stimulation (TUS) key parameters. Shown are the abbreviations and measurement value definitions for TUS parameters.

| PARAMETERS | DESCRIPTION | DEFINITION | UNITS |
| --- | --- | --- | --- |
| **ISPPA** | Intensity - Spatial Peak<br>Pulse Average | Refers to the Average acoustic intensity | W/cm<sup>2</sup> |
| **ISPTA** | Intensity - Spatial Peak<br>Temporal Average | Temporally averaged intensity over the<br>sonication duration (ISPPA * Duty Cycle) | W/cm<sup>2</sup> |
| **SD** | Sonication Duration | Period during which TUS is applied to<br>the brain target | Seconds |
| **DC** | Duty Cycle | Ratio of sonication on and off time | % |
| **PRF** | Pulse Repetition<br>Frequency | The number of pulses per second<br>delivered to the target | Hz |
| **MI** | Mechanical Index | Characterizes the likelihood of<br>mechanical cavitation caused by TUS |  |
| **TI** | Thermal Index | Estimate of temperature rise in tissue<br>due to TUS exposure. |  |

**Box 2.** TUS recommendations. The International Transcranial Ultrasonic Stimulation Safety and Standards (ITRUSST) consortium has recently established recommendations based on existing guidelines for diagnostic ultrasound from regulatory bodies such as the Food and Drug Administration (FDA), the British Medical Ultrasound Society (BMUS) and the American Institute of Ultrasound in Medicine (AIUM). Some in the community might follow the FDA guidelines for diagnostic ultrasound, see Box 1 for a description of these parameters. The ITRUSST consortium concurs with the FDA limits on MI, but differs on the importance placed on the intensity (ISPPA/ISPTA) parameters independently of consideration of the TI (noted by * in the box above). Namely, ITRUSST recommends that the thermal maximal exposure is ≤ 2° Celsius at any time, the thermal dose is ≤ 0.25 CEM43 or the maximum exposure time is also considered (e.g., 10 sec 5.0 < TI ≤ 6.0), please see (Aubry et al., 2023). Importantly, the ITRUSST recommendations should be considered in parallel to individualized simulations and measurement of thermal effects to further reduce the risk of adverse effects.

LIMITS FOR DIAGNOSTIC ULTRASOUND
|  |  |
| --- | --- |
| ISPPA | $\leq 190 \text{ W/cm}^2$ |
| ISPTA | $\leq 720 \text{ mW/cm}^2*$ |
| MI | $\leq 1.9$ |
| TI | $< 6^*$ |

### Directionality of TUS Hypotheses

**Box 3.** Net enhancement versus suppression hypotheses. Summarized hypotheses on how net enhancement or suppression could be biased with TUS parameters. Note that the threshold for ISPPA qualifying as ‘higher’ or ‘lower’ intensity is currently poorly understood or may non-linearly interact with other factors such as Duty Cycle (Plaksin et al., 2016; Plaksin et al., 2014). A similar set of uncertainty in the directionality and border exists for the other parameters, here defined as falsifiable hypotheses.

|  | Intensity (ISPPA brain) | Sonation duration | Duty Cycle (DC) | Pulse Repetition Frequency (PRF) |
| --- | --- | --- | --- | --- |
| Enhancement weighted | Higher intensity ($> 1 \text{ W/cm}^2$) | $< 500 \text{ ms}$ | $> 30\%$ | $> 300 \text{ Hz}$ |
| Suppression weighted | Lower intensity ($< 1 \text{ W/cm}^2$) | $> 500 \text{ ms}$ | $< 30\%$ | $< 300 \text{ Hz}$ |

## Introduction to the Iowa-Newcastle (inTUS)

### Transcranial Ultrasound Stimulation Resource

The inTUS Resource consists of the following resource items, linked to Caffaratti et al. ***Neuromodulation with Ultrasound: Hypotheses on the Directionality of Effects and a Community Resource.*** Please cite the latest eLife paper when using and referring to the inTUS resource: https://elifesciences.org/reviewed-preprints/100827.

The inTUS Resource documents can be found on the Laboratory of Comparative Neuropsychology data share on the Open Science Framework: https://osf.io/arqp8/ under the folder Cafferatti_et_al_inTUS_Resource. If the location of the repository changes, a link to its location will be left here.

#### Resource documents

- **Database Tables**

◦ inTUS_DATABASE_1-2025.csv

▪ Certain key historic versions will be retained, e.g., the initial inTUS_DATABASE_6-2024.csv
◦ <u>InTUS_DATABASE_1-2025_description.pdf</u> describes the columns in the database
- **R Markdown script** to regenerate the figures and statistics using R Studio, and example PDF/HTML output if not using R to more quickly visualize TUS parameter effects.

◦ <u>TUS_Effects_1-2025_Ranalyses.Rmd</u> is the executable R markdown script, which executes the statistical models and figures. To run the script, install the freely available R Studio software (https://posit.co/). Run the script sections as described in the instructions at the top of the script (tested in R Studio version RStudio-2024.12.0-467).
◦ <u>InTUS_OUTPUT_1-2025.pdf</u> is an example output file generated by the latest database, if the reader more quickly wants to see the latest output without having to run the R markdown script: https://rpubs.com/BenSlaterNeuro/1268823
◦ <u>Data visualization online</u> app to visualize the TUS parameters data: https://benslaterneuro.shinyapps.io/Caffaratti_inTUS_Resource/
- **To contribute to the database please use the Qualtrics Survey**

◦ **Qualtrics Survey** to submit your paper and data so that it can be considered for addition to the database. Please use this link: https://uiowa.qualtrics.com/jfe/form/SV_4VOvb0fdwvACDkO

#### Using the resource

To regenerate the manuscript figures with the latest tables, Download the latest *database table* and *R_script*. Run the script in R or R Studio making sure it can access the inTUS database you are interested in (e.g., that the database is visible in the R Studio working directory).

#### Contributing to the resource

Please use the Qualtrics Link and Form to submit your preprint or published paper data. These submissions will be checked and once verified will be added to the database: https://uiowa.qualtrics.com/jfe/form/SV_4VOvb0fdwvACDkO

#### How the resource is curated

To ensure that the database has TUS researcher checked information, we have established the Qualtrics form that can be used to submit a preprint or publication citation and associated TUS values and outcomes. The human TUS expert factor ensures that a skilled researcher checks the submitted data that could be included in the database. Please note the inclusionary and exclusionary criteria in the paper, which may change with future database expansions. We are exploring the possibility of a collaboration with the ITRUSST consortium to sustain the resource and will update the OSF link if the location of the inTUS resource changes.

#### Only healthy humans?

This resource was initially established with healthy human participant low-intensity TUS studies. We will continue to expand the inTUS resource with the TUS community’s help and input. We are open to the resource being extended to human patients and nonhuman animals of different species. Please use the Qualtrics Form if you’re interested in helping or have suggestions about the resource.

## Acknowledgements

Supported by National Institutes of Health USA (R01–DC04290; BRAIN Initiative U01-NS137991), National Science Foundation (SBE-UKRI-2342847), and Medical Research Council (UK). Ben Slater is supported by a BBSRC UK PhD studentship. M.Ka. was supported by the Engineering and Physical Sciences Research Council (EP/W004488/1 and EP/X01925X/1) and the Guangci Professorship Program of Rui Jin Hospital (Shanghai Jiao Tong University)

## Data availability

The datasets and R script generated in this study have been deposited in the Open Science Framework https://osf.io/arqp8/ in the ‘Caffaratti_et_al_inTUS_Resource’ folder.

